# Meta-analysis of 208,370 East Asians identifies 113 genomic loci and yields new non-immune cell relevant biological insights for systemic lupus erythematosus

**DOI:** 10.1101/2020.08.22.20178939

**Authors:** Xianyong Yin, Kwangwoo Kim, Hiroyuki Suetsugu, So-Young Bang, Leilei Wen, Masaru Koido, Eunji Ha, Lu Liu, Yuma Sakamoto, Sungsin Jo, Rui-Xue Leng, Nao Otomo, Viktoryia Laurynenka, Young-Chang Kwon, Yujun Sheng, Nobuhiko Sugano, Mi Yeong Hwang, Weiran Li, Masaya Mukai, Kyungheon Yoon, Minglong Cai, Kazuyoshi Ishigaki, Won Tae Chung, He Huang, Daisuke Takahashi, Shin-Seok Lee, Mengwei Wang, Kohei Karino, Seung-Cheol Shim, Xiaodong Zheng, Tomoya Miyamura, Young Mo Kang, Dongqing Ye, Junichi Nakamura, Chang-Hee Suh, Yuanjia Tang, Goro Motomura, Yong-Beom Park, Huihua Ding, Takeshi Kuroda, Jung-Yoon Choe, Chengxu Li, Hiroaki Niiro, Youngho Park, Changbing Shen, Takeshi Miyamoto, Ga-Young Ahn, Wenmin Fei, Tsutomu Takeuchi, Jung-Min Shin, Keke Li, Yasushi Kawaguchi, Yeon-Kyung Lee, Yongfei Wang, Koichi Amano, Wanling Yang, Yoshifumi Tada, Ken Yamaji, Masato Shimizu, Takashi Atsumi, Akari Suzuki, Takayuki Sumida, Yukinori Okada, Koichi Matsuda, Keitaro Matsuo, Yuta Kochi, Japanese Research Committee on Idiopathic Osteonecrosis of the Femoral Head, Leah C. Kottyan, Matthew T. Weirauch, Sreeja Parameswaran, Shruti Eswar, Hanan Salim, Xiaoting Chen, Kazuhiko Yamamoto, John B. Harley, Koichiro Ohmura, Tae-Hwan Kim, Sen Yang, Takuaki Yamamoto, Bong-Jo Kim, Nan Shen, Shiro Ikegawa, Hye-Soon Lee, Xuejun Zhang, Chikashi Terao, Yong Cui, Sang-Cheol Bae

## Abstract

Systemic lupus erythematosus (SLE), an autoimmune disorder, has been associated with nearly 100 susceptibility loci^1-8^. Nevertheless, these loci only partially explain SLE heritability and provide limited biological insight. We report the largest study of SLE in East Asians (13,377 cases and 194,993 controls), identifying 233 association signals within 113 (46 novel) genetic loci. We detect six new lead missense variants and prioritize ten most likely putative causal variants, one of which we demonstrate exhibits allele-specific regulatory effect on *ACAP1* in vitro. We suggest 677 effector genes with potential for drug repurposing, and provide evidence that two distinct association signals at a single locus act on different genes (*NCF2* and *SMG7)*. We demonstrate that SLE-risk variants overlap with cell-specific active regulatory elements, notably EBNA2-mediated super-enhancers in Epstein-Barr Virus-transformed B cells, and implicate the role for non-immune cells in SLE biology. These findings shed light on genetic and biological understandings of SLE.

## Main text

Systemic lupus erythematosus (SLE) is an autoimmune disorder characterized by the production of autoantibodies that damage multiple organs^9^. Considerable genetic predisposition contributes to SLE etiology^10^. To date, nearly 100 susceptibility loci have been identified for SLE, mainly through genome-wide association studies (GWASs)^1-4,7,8^. However, these loci collectively only explain ~30% of SLE heritability^5^ and their biology, in terms of causal variants, effector genes and cell types, and pathological pathways that mediate genetic effects, has not yet been fully characterized^11^.

Genome-wide association meta-analyses (GWMA) have been performed to uncover new genetic associations for SLE in Asians^6^, Europeans^12^, and trans-ancestral populations^5^. However, the study sample sizes were relatively modest, which limits their ability for genetic discovery. GWASs have successfully linked genetic variants with human common diseases and traits^13^. Nonetheless, only ~8% of GWAS participants are East Asians^14^. East Asians have a unique population genetic history and may have unique genetic disease risk mechanisms and exhibit specific disease manifestations. For example, SLE has a remarkably higher prevalence and younger age of onset in Asians^15,16^. Genetic heterogeneity may explain, at least partly, the phenotypic diversity of SLE between East Asians and Europeans^5^. Hence, large-scale East Asian investigations may identify unique genetic associations even for the same diseases and traits that have already been well studied in Europeans^17^.

Here, we report the largest-ever GWMA for SLE in 208,370 East Asians. Our study identified many new genetic associations, some of which have not been detected in Europeans, and improved our biological understanding of SLE pathophysiology.

We newly generated three GWAS datasets from 10,029 SLE cases and 180,167 controls, which we meta-analyzed together with five published studies (3,348 cases and 14,826 controls)^1,2,4,8,18^ after whole-genome genotype imputation using 1000 Genomes Project phase 3^19^ and population-specific reference panels^20^, bringing the total sample size to 208,370 **(Supplementary Table 1)**. To the best of our knowledge, this is the largest genetic association study of SLE to date. The effective sample size (N_eff_=50,072) is three- and four-fold larger than that of the largest published trans-ancestry^5^ and East Asian^6^ meta-analyses, respectively.

We tested associations for 11,270,530 genetic variants in a fixed-effects meta-analysis. A quantile-quantile plot showed that test statistics were well-calibrated, with a genomic-control inflation factor A_GC_=1-06 (indicating that ancestry effects had been well controlled; **Supplementary Figure 1)**. Linkage disequilibrium (LD) score regression^21^ showed that polygenic effects (89.4%), rather than biases, primarily caused the inflation residual (estimated mean χ^2^=1.32 and LD-score intercept=1.03).

We detected 26,379 genetic variants associated with SLE at P<5×10^-8^ within 113 loci **(Supplementary Table 2** and **Fig. 1a)**, of which 46 were novel **(Tab. 1)**. The pairwise LD between lead variants is low (LD r^2^<0.002). For seven novel loci, minor allele frequencies (MAFs) of the lead single nucleotide polymorphisms (SNPs) were ten-fold higher in East Asians than in Europeans **(Fig. 1b)**. Two of them and their neighbors in strong LD (LD r^2^≥0.2 in either East Asians or Europeans) would be undetectable in Europeans at the same effective sample size and risk magnitude as we find in East Asians (statistical power<10%; **Supplementary Table 3)**.

**Table 1:**
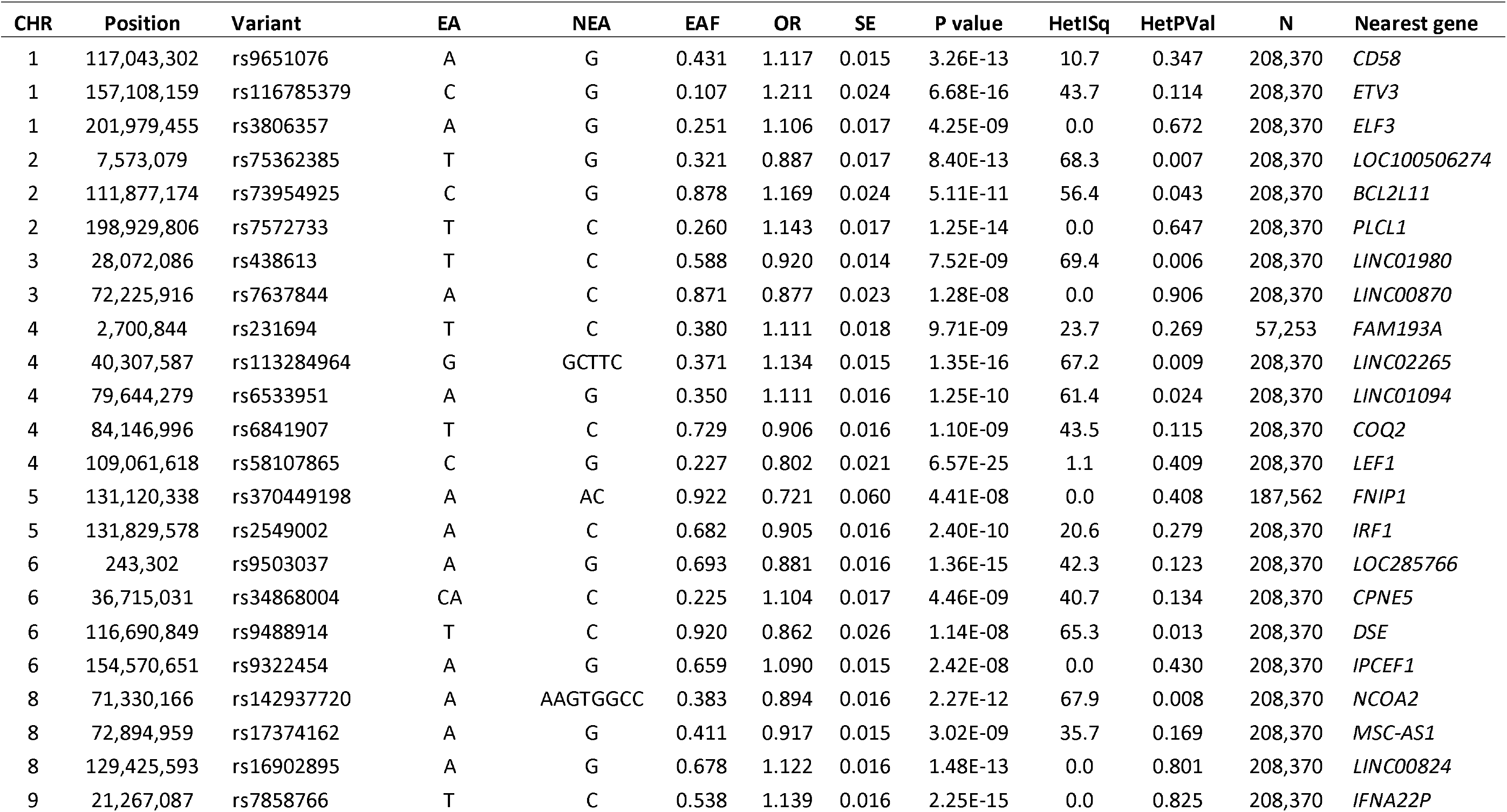
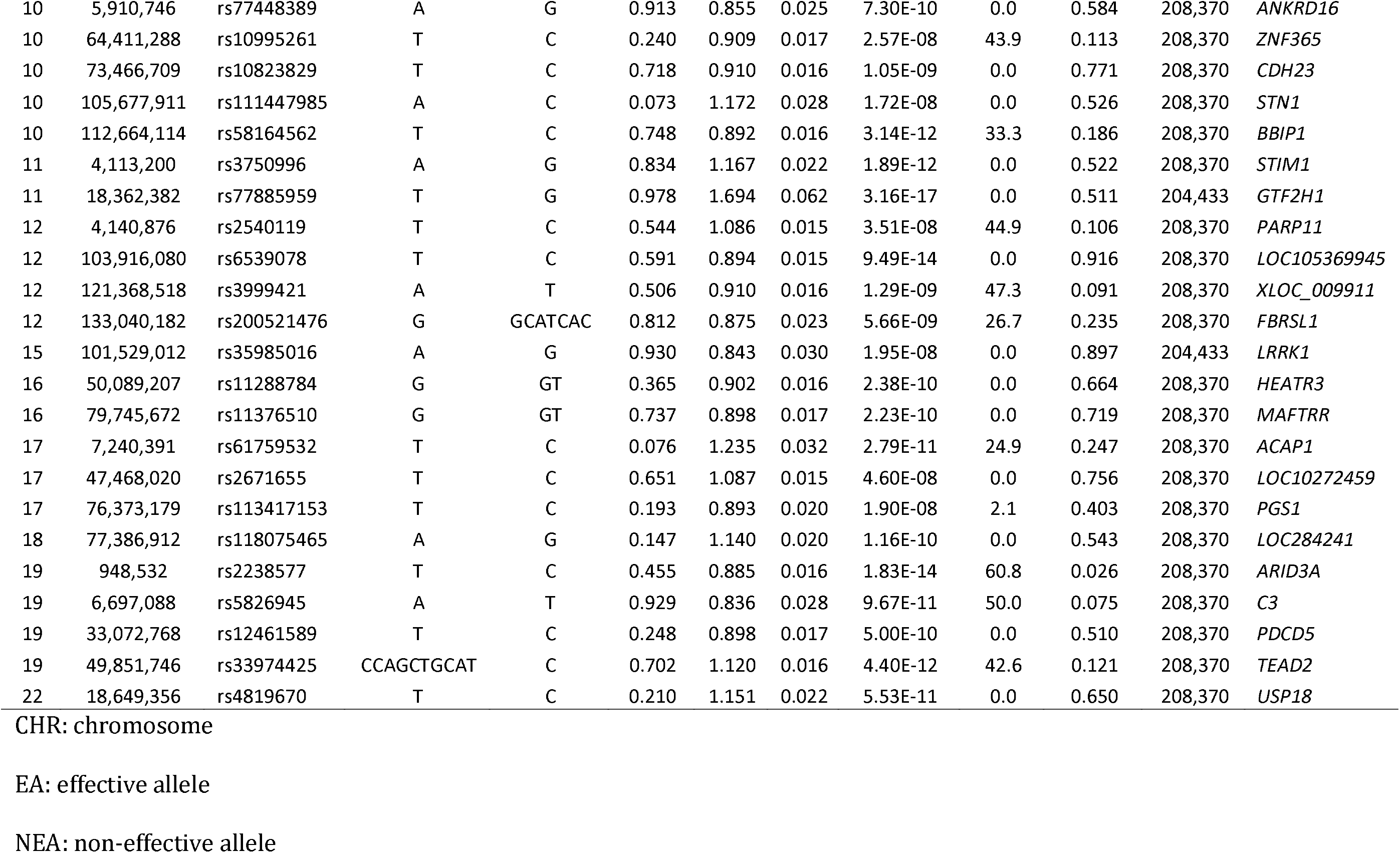
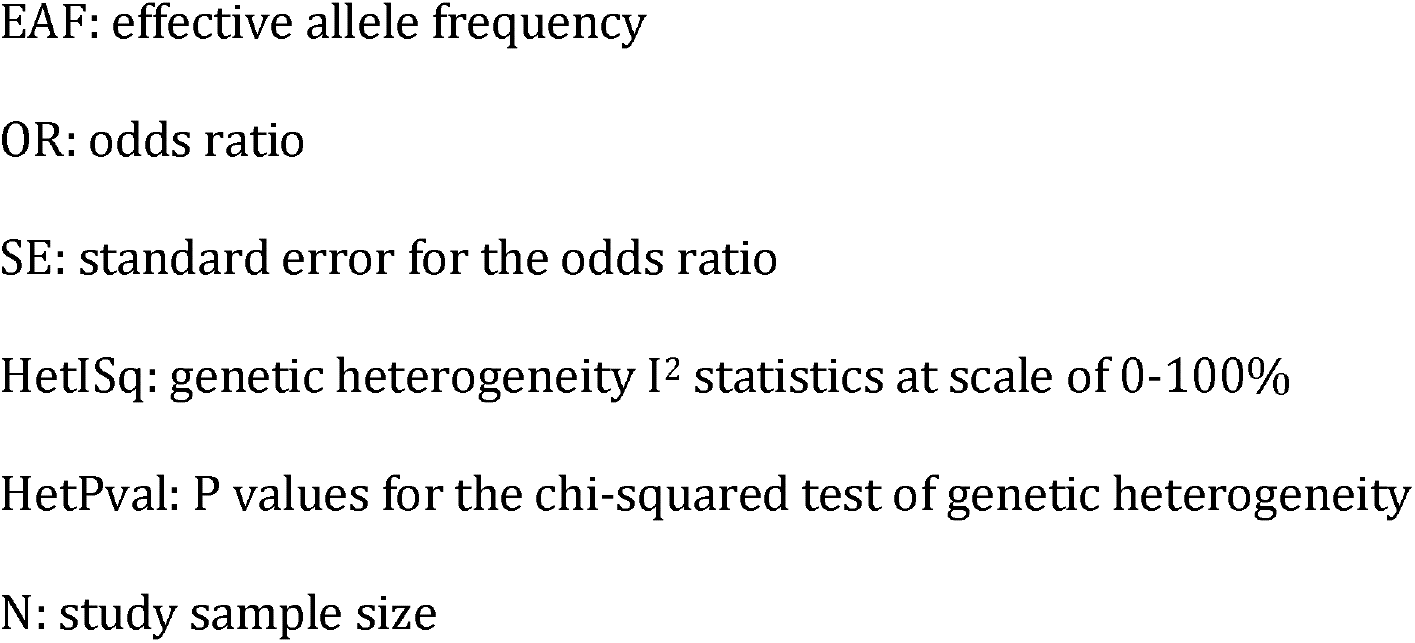
Association results for the 46 novel susceptibility loci for SLE.

| CHR | Position | Variant | EA | NEA | EAF | OR | SE | P value | HetISq | HetPVal | N | Nearest gene |
| --- | --- | --- | --- | --- | --- | --- | --- | --- | --- | --- | --- | --- |
| 1 | 117,043,302 | rs9651076 | A | G | 0.431 | 1.117 | 0.015 | 3.26E-13 | 10.7 | 0.347 | 208,370 | *CD58* |
| 1 | 157,108,159 | rs116785379 | C | G | 0.107 | 1.211 | 0.024 | 6.68E-16 | 43.7 | 0.114 | 208,370 | *ETV3* |
| 1 | 201,979,455 | rs3806357 | A | G | 0.251 | 1.106 | 0.017 | 4.25E-09 | 0.0 | 0.672 | 208,370 | *ELF3* |
| 2 | 7,573,079 | rs75362385 | T | G | 0.321 | 0.887 | 0.017 | 8.40E-13 | 68.3 | 0.007 | 208,370 | *LOC100506274* |
| 2 | 111,877,174 | rs73954925 | C | G | 0.878 | 1.169 | 0.024 | 5.11E-11 | 56.4 | 0.043 | 208,370 | *BCL2L11* |
| 2 | 198,929,806 | rs7572733 | T | C | 0.260 | 1.143 | 0.017 | 1.25E-14 | 0.0 | 0.647 | 208,370 | *PLCL1* |
| 3 | 28,072,086 | rs438613 | T | C | 0.588 | 0.920 | 0.014 | 7.52E-09 | 69.4 | 0.006 | 208,370 | *LINC01980* |
| 3 | 72,225,916 | rs7637844 | A | C | 0.871 | 0.877 | 0.023 | 1.28E-08 | 0.0 | 0.906 | 208,370 | *LINC00870* |
| 4 | 2,700,844 | rs231694 | T | C | 0.380 | 1.111 | 0.018 | 9.71E-09 | 23.7 | 0.269 | 57,253 | *FAM193A* |
| 4 | 40,307,587 | rs113284964 | G | GCTTC | 0.371 | 1.134 | 0.015 | 1.35E-16 | 67.2 | 0.009 | 208,370 | *LINC02265* |
| 4 | 79,644,279 | rs6533951 | A | G | 0.350 | 1.111 | 0.016 | 1.25E-10 | 61.4 | 0.024 | 208,370 | *LINC01094* |
| 4 | 84,146,996 | rs6841907 | T | C | 0.729 | 0.906 | 0.016 | 1.10E-09 | 43.5 | 0.115 | 208,370 | *COQ2* |
| 4 | 109,061,618 | rs58107865 | C | G | 0.227 | 0.802 | 0.021 | 6.57E-25 | 1.1 | 0.409 | 208,370 | *LEF1* |
| 5 | 131,120,338 | rs370449198 | A | AC | 0.922 | 0.721 | 0.060 | 4.41E-08 | 0.0 | 0.408 | 187,562 | *FNIP1* |
| 5 | 131,829,578 | rs2549002 | A | C | 0.682 | 0.905 | 0.016 | 2.40E-10 | 20.6 | 0.279 | 208,370 | *IRF1* |
| 6 | 243,302 | rs9503037 | A | G | 0.693 | 0.881 | 0.016 | 1.36E-15 | 42.3 | 0.123 | 208,370 | *LOC285766* |
| 6 | 36,715,031 | rs34868004 | CA | C | 0.225 | 1.104 | 0.017 | 4.46E-09 | 40.7 | 0.134 | 208,370 | *CPNE5* |
| 6 | 116,690,849 | rs9488914 | T | C | 0.920 | 0.862 | 0.026 | 1.14E-08 | 65.3 | 0.013 | 208,370 | *DSE* |
| 6 | 154,570,651 | rs9322454 | A | G | 0.659 | 1.090 | 0.015 | 2.42E-08 | 0.0 | 0.430 | 208,370 | *IPCEF1* |
| 8 | 71,330,166 | rs142937720 | A | AAGTGGCC | 0.383 | 0.894 | 0.016 | 2.27E-12 | 67.9 | 0.008 | 208,370 | *NCOA2* |
| 8 | 72,894,959 | rs17374162 | A | G | 0.411 | 0.917 | 0.015 | 3.02E-09 | 35.7 | 0.169 | 208,370 | *MSC-AS1* |
| 8 | 129,425,593 | rs16902895 | A | G | 0.678 | 1.122 | 0.016 | 1.48E-13 | 0.0 | 0.801 | 208,370 | *LINC00824* |
| 9 | 21,267,087 | rs7858766 | T | C | 0.538 | 1.139 | 0.016 | 2.25E-15 | 0.0 | 0.825 | 208,370 | *IFNA22P* |
| 10 | 5,910,746 | rs77448389 | A | G | 0.913 | 0.855 | 0.025 | 7.30E-10 | 0.0 | 0.584 | 208,370 | ANKRD16 |
| 10 | 64,411,288 | rs10995261 | T | C | 0.240 | 0.909 | 0.017 | 2.57E-08 | 43.9 | 0.113 | 208,370 | ZNF365 |
| 10 | 73,466,709 | rs10823829 | T | C | 0.718 | 0.910 | 0.016 | 1.05E-09 | 0.0 | 0.771 | 208,370 | CDH23 |
| 10 | 105,677,911 | rs111447985 | A | C | 0.073 | 1.172 | 0.028 | 1.72E-08 | 0.0 | 0.526 | 208,370 | STN1 |
| 10 | 112,664,114 | rs58164562 | T | C | 0.748 | 0.892 | 0.016 | 3.14E-12 | 33.3 | 0.186 | 208,370 | BBIP1 |
| 11 | 4,113,200 | rs3750996 | A | G | 0.834 | 1.167 | 0.022 | 1.89E-12 | 0.0 | 0.522 | 208,370 | STIM1 |
| 11 | 18,362,382 | rs77885959 | T | G | 0.978 | 1.694 | 0.062 | 3.16E-17 | 0.0 | 0.511 | 204,433 | GTF2H1 |
| 12 | 4,140,876 | rs2540119 | T | C | 0.544 | 1.086 | 0.015 | 3.51E-08 | 44.9 | 0.106 | 208,370 | PARP11 |
| 12 | 103,916,080 | rs6539078 | T | C | 0.591 | 0.894 | 0.015 | 9.49E-14 | 0.0 | 0.916 | 208,370 | LOC105369945 |
| 12 | 121,368,518 | rs3999421 | A | T | 0.506 | 0.910 | 0.016 | 1.29E-09 | 47.3 | 0.091 | 208,370 | XLOC_009911 |
| 12 | 133,040,182 | rs200521476 | G | GCATCAC | 0.812 | 0.875 | 0.023 | 5.66E-09 | 26.7 | 0.235 | 208,370 | FBRSL1 |
| 15 | 101,529,012 | rs35985016 | A | G | 0.930 | 0.843 | 0.030 | 1.95E-08 | 0.0 | 0.897 | 204,433 | LRRK1 |
| 16 | 50,089,207 | rs11288784 | G | GT | 0.365 | 0.902 | 0.016 | 2.38E-10 | 0.0 | 0.664 | 208,370 | HEATR3 |
| 16 | 79,745,672 | rs11376510 | G | GT | 0.737 | 0.898 | 0.017 | 2.23E-10 | 0.0 | 0.719 | 208,370 | MAFTRR |
| 17 | 7,240,391 | rs61759532 | T | C | 0.076 | 1.235 | 0.032 | 2.79E-11 | 24.9 | 0.247 | 208,370 | ACAP1 |
| 17 | 47,468,020 | rs2671655 | T | C | 0.651 | 1.087 | 0.015 | 4.60E-08 | 0.0 | 0.756 | 208,370 | LOC10272459 |
| 17 | 76,373,179 | rs113417153 | T | C | 0.193 | 0.893 | 0.020 | 1.90E-08 | 2.1 | 0.403 | 208,370 | PGS1 |
| 18 | 77,386,912 | rs118075465 | A | G | 0.147 | 1.140 | 0.020 | 1.16E-10 | 0.0 | 0.543 | 208,370 | LOC284241 |
| 19 | 948,532 | rs2238577 | T | C | 0.455 | 0.885 | 0.016 | 1.83E-14 | 60.8 | 0.026 | 208,370 | ARID3A |
| 19 | 6,697,088 | rs5826945 | A | T | 0.929 | 0.836 | 0.028 | 9.67E-11 | 50.0 | 0.075 | 208,370 | C3 |
| 19 | 33,072,768 | rs12461589 | T | C | 0.248 | 0.898 | 0.017 | 5.00E-10 | 0.0 | 0.510 | 208,370 | PDCD5 |
| 19 | 49,851,746 | rs33974425 | CCAGCTGCAT | C | 0.702 | 1.120 | 0.016 | 4.40E-12 | 42.6 | 0.121 | 208,370 | TEAD2 |
| 22 | 18,649,356 | rs4819670 | T | C | 0.210 | 1.151 | 0.022 | 5.53E-11 | 0.0 | 0.650 | 208,370 | USP18 |

CHR: chromosome
EA: effective allele
NEA: non-effective allele
EAF: effective allele frequency
OR: odds ratio
SE: standard error for the odds ratio
HetISq: genetic heterogeneity $I^2$ statistics at scale of 0-100%
HetPval: P values for the chi-squared test of genetic heterogeneity
N: study sample size

**Fig. 1.**
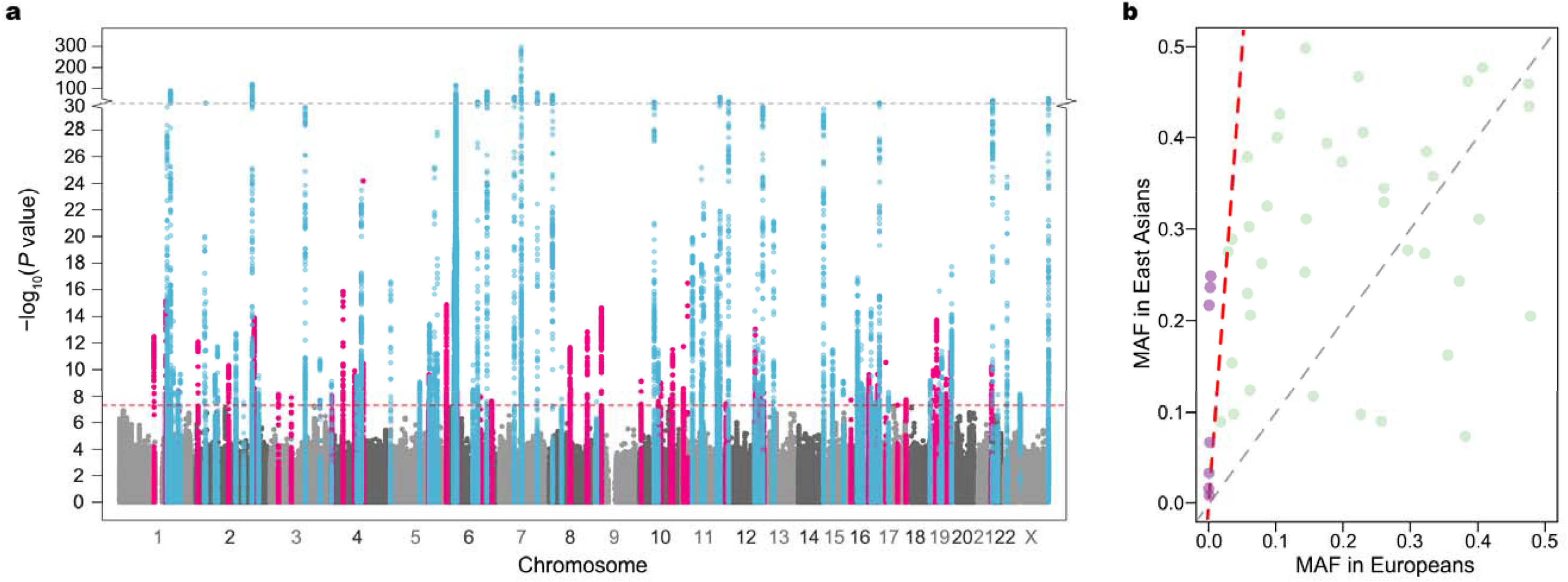
Summary of meta-analysis association results and comparison of MAFs for lead variants within the 46 novel loci between East Asians and Europeans, **a**, Manhattan plot of genome-wide association meta-analysis results from 208,370 SLE case-control East Asians. Minus logio-transformed association *P* values (y-axis) are plotted along chromosomal positions (x-axis). Known and novel loci are highlighted in light blue and pink, respectively. The red dashed line denotes the genome-wide association significance threshold of *P=* 5×10^-8^. The gray dashed line represents *P*=10~^30^, at which the y-axis breaks, **b**, Comparison of MAFs of lead variants within the 46 novel loci between East Asians (y-axis) and non-Finnish Europeans (x-axis) in the Genome Aggregation Database (gnomAD) version 3. Variants with more than ten-times higher MAFs in East Asians are colored purple above a red dashed line.

To dissect the source of association signals at each locus, we conducted an approximate conditional analysis using GCTA^22^ with meta-analysis summary statistics and LD estimates from 7,021 unrelated Chinese controls **(Online Methods)**. We acknowledge the limitations of using LD estimation from a single population for a meta-analysis of diverse East Asians. We identified a total of 233 independent association signals with conditional P<5×10^-8^,169 of which arose from non-Human Leukocyte Antigen (*HLA)* regions **(Supplementary Table 4)**. We observed two to four signals at each of 28 non*-HLA* loci (including 7 novel loci). For example, we discovered two distinct association signals within the known *STAT4* locus, including the previously reported SNPrs11889341^12^ and the new insert-deletion variant (indel) rs71403211 **(Extended Data Fig. 1a)**. For the 46 novel loci, we discovered 55 distinct signals **(Supplementary Table 4** and **Supplementary Figure 2)**. Most of the signal index variants (n=190, 82%) are common (MAF≥5%) with modest effects **(Supplementary Table 4)**.

We identified 11 exonic signal index variants at ten non*-HLA* loci **(Supplementary Table 4)**, highlighting the roles of *AHNAK2, CSK, IKBKB, IRAK1, NCF2, OAS1, TYK2*, and *WDFY4* within eight known loci, and *CDH23* and *LRRK1* within two novel loci **(Extended Data Fig. 1-2)**. We detected two distinct signals within *WDFY4*, including the known (rs7097397)^18^ and a new (rs7072606) missense variant (LD r^2^=0.02 in East Asians), which suggest a potential allelic series effect at this locus **(Extended Data Fig. 1b)**. We replicated the association of the missense variants *at AHNAK2* (rs2819426)^23^, *IRAK1* (rs1059702)^24^, and *NCF2* (rs13 3 0 6 5 75)^25,26^, and provided for the first time genome-wide association evidence at a missense variant within *OAS1* (rs1131476, LD r^2^=0.78 with the known missense variant rs1051042 that attains suggestive significance in East Asians)^23^. We detected three new exonic variants (including two missense variants) within the *CSK*(rs11553760), *1KBKB* (rs2272736), and *TYK2* (rs55882956) genes **(Extended Data Fig. 2)**. They were not correlated with previously reported exonic variants within the same genes (LD r^2^<0.02 in East Asians or Europeans; **Supplementary Table 5)**, suggesting possible allelic heterogeneity of these genes. In the two novel loci with lead missense variants **(Extended Data Fig. 2)**, *CHD23* plays a role in cell migration^27^ while *LRRK1* encodes a multiple-domain leucine-rich repeat kinase. A previous study observed that *LRRK1*-deficient mice exhibited a profound defect in B-cell proliferation and survival and impaired B-cell receptor-mediated NF-κB activation^28^.

To prioritize putative causal variants, we conducted a Bayesian statistical fine-mapping analysis for 111 loci using FINEMAP^29^ after excluding complex associations involving *HLA* and **7q11.23**. We found exactly the same number of association signals in **57** loci between FINEMAP causal configuration with the highest posterior probability (PP) and the GCTA approximate conditional test. To be conservative, we only summarized the statistical fine-mapping results for these 57 regions, which contained **65** association signals **(Supplementary Table 6)**.

For each signal, we built a credible set of putative causal variants with a 95% probability of including the true causal variants. The size of 28 credible sets was small (size≤10; **Fig. 2a)**. Among the 110 putative causal variants with PP≥0.1 **(Fig. 2b)**, we found four coding variants (3.6%), which implies that most of these associations are probably induced by non-coding causal variants. The prioritized variants are available to be tested as potential targets in perturbation experiments. For example, the allele-specific regulatory activity of the intronic variant (rs 10036748) with the highest PP (0.387) in the *TNIP1* locus was recently characterized in SLE^30^.

**Fig. 2.**
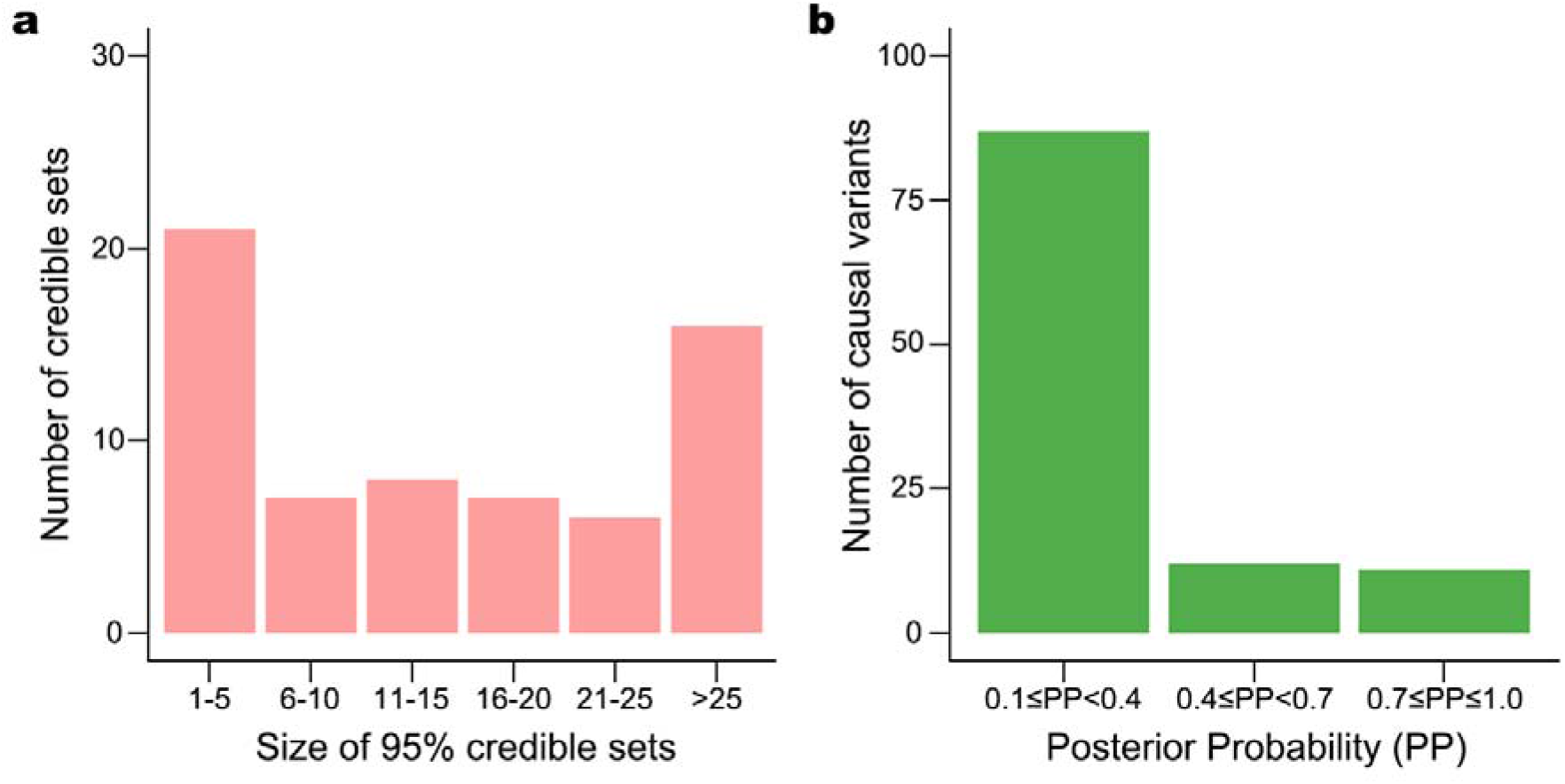
Results of statistical fine-mapping. **a**, Number of 95% credible sets of putative causal variants, binned by their sizes, **b**, Number of potential causal variants with posterior probabilities (PP) ≥ 0.1, which are considered to be the true causal variants.

We pinpointed a single most likely causal variant with high confidence (PP≥0.8) for four known (*ATXN2, BACH2, DRAM1/WASHC3*, and *NCF2*) and six novel (17p13.1, *ELF3, GTF2H1, LRRK1, WC102724596/PHB*, and *ST1M1*) loci **(Supplementary Table 6)**. For example, we prioritized rs61759532 as a putative causal variant at the novel 17p13.1 locus (PP=0.999; **Fig. 3a)**. This variant is located in an intron of *ACAP1*, which encodes a key regulator of integrin traffic for cell adhesion and migration^31^. We observed that rs61759532 overlaps with an accessible open chromatin region in blood B and T cells **(Fig. 3b)**. Transcriptional reporter assays showed significant allelic differences in the enhancer activity of rs61759532 in THP1 monocyte cell lines (two-sided t-test P=8.1×10^-3^; **Fig. 3c)**, consistent with the regulatory effect of the risk allele, *T*, in reducing *ACAP1* expression levels in whole blood^32^(P=1.7×10^-47^; **Fig. 3d)**. Electrophoretic mobility shift assays (EMSA) revealed that allele-specific biotin-labeled probes containing the *T* (risk allele) form fewer nuclear protein-probe complexes than probes with *C* (non-risk allele) in THP1 and Epstein-Barr virus (EBV)-transformed B cell lines **(Fig. 3e)**.

**Fig. 3.**
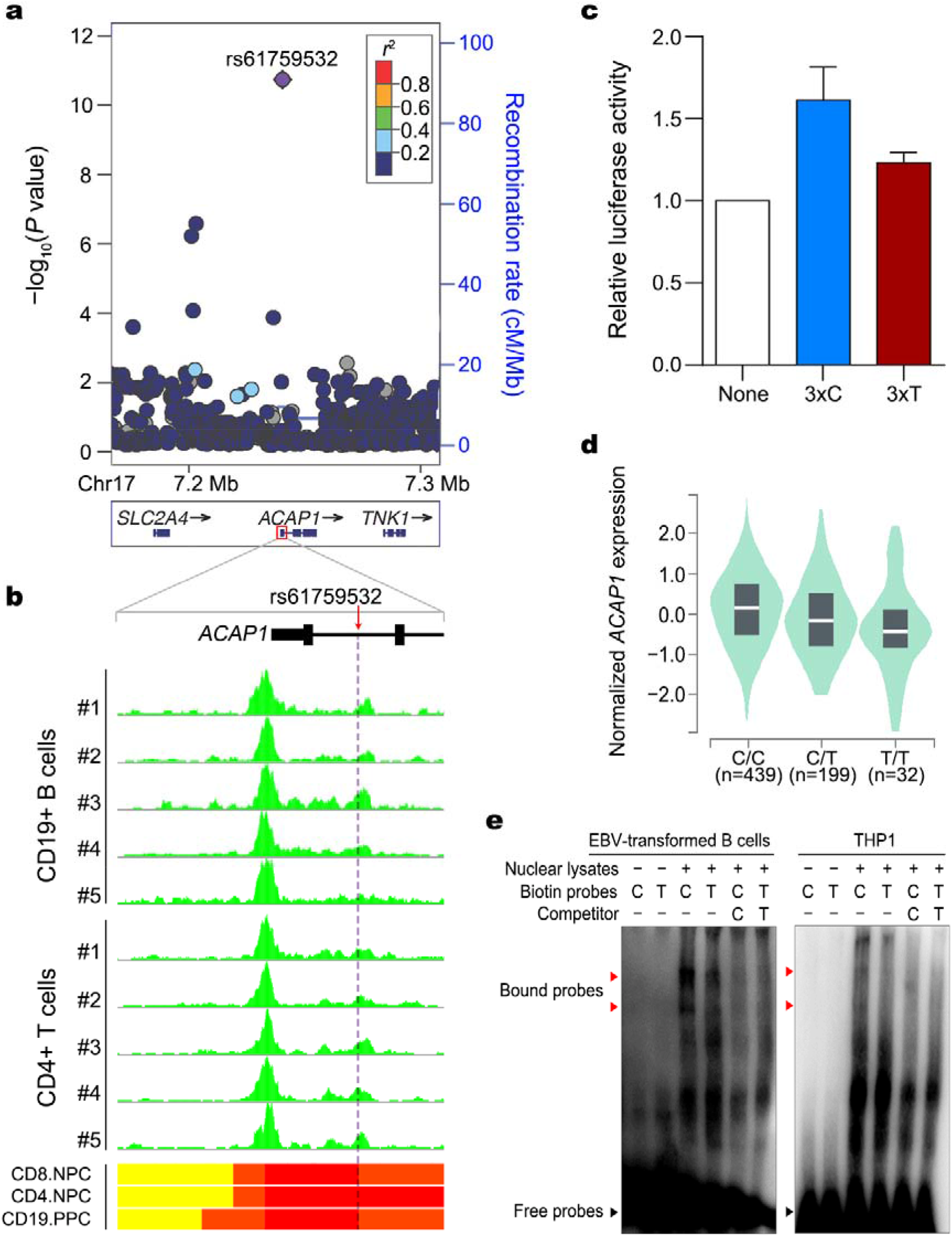
Allele-specific regulatory effect of rs61759532 on *ACAP1*. **a**, Regional association plot for the *ACAP1* locus. The lead variant rs61759532 is labeled as a purple diamond. The LD was estimated using data from 7,021 Chinese individuals, **b**, Location of rs61759532 within an ATAC-seq open chromatin accessible region in CD19^+^ B- and CD4^+^ T-cells (green tracks) and within active ChromHMM chromatin states (bars on the bottom panel) in primary CD8^+^ T naive cells (CD8.NPC), T helper naive cells (CD4.NPC), and primary B cells (BLD.CD19.PPC). Chromatin states are colored red (active transcription start site), orange red (flanking active transcription start site, and yellow (enhancers), **c**, Allelic differentials in the enhancing activity of rs61759532 in THP1 cells. None, 3×C, and 3×T denote empty vector containing a minimal promoter, and vectors with the *C* and *T* allele of rs61759532, respectively. Relative luciferases activities, measured in five independent biological replicates, were significantly higher for inserts with the *C* allele (P=8.1×10^-3^; two-tailed t-test). Error bars indicate standard errors of the means of five independent biological replicates, **d**, Association between the risk allele (7) of rs61759532 and decreased expression of *ACAP1* in GTEx v8 whole blood (P=1.7×10^-47^). The white line in the center of each box indicates median expression value, while the box for each genotype represents the interquartile range of *ACACP1* expressions, **e**, Allelic differential in protein-DNA binding by rs61759532 in EMSAs. Biotin-conjugated 30-nucleotide probes flanking rs61759532 (denoted as *C* or *T*, according the allele) were incubated with nuclear extracts (10 μg) from EBV-transformed B cells or THP1 cells in EMSAs. Shifted bands (indicated by red arrows) had stronger intensities with biotin-conjugated *C* allele probes than *T* allele probes, and were not detected in the presence of excess non-conjugated probes.

We deployed four gene-level methods to comprehensively catalog potential effector genes. Briefly, we performed a transcriptome-wide association study (TWAS)^33^ using expression quantitative trait loci (eQTL) from six human immune cell types in up to 105 East Asians^34^, a gene-based association analysis using Multi-marker Analysis of GenoMic Annotation (MAGMA)^35^, and two additional data-mining approaches^36,37^ that integrated genetic associations with gene function and chromosomal position, eQTL, and chromatin interactions **(Online Methods)**. We nominated 677 possible effector genes **(Supplementary Table 7-11** and **Extended Data Fig. 3)**. Of the 677 genes, 285 (42%) were supported by at least two approaches and 222 (35.4%) were predicted to have differential expression levels in SLE in blood immune cells from East Asian individuals, including 26 genes from regions (<500 kb) where genome-wide significant associations have not previously been reported. For example, we found significant evidence for the *FAS* gene only by TWAS. This gene encodes a member of the TNF receptor superfamily and plays a central role in lymphocyte apoptosis. Individuals with defects in the Fas/FasL system develop lupus-like symptoms^38^.

The gene-level analysis provided an opportunity to interrogate the biology of SLE loci. For example, we detected two independent association signals within the known SLE locus including *SMG7* and *NCF2* **(Fig. 4a)**. The lead SNP rs13306575 is a missense variant, which substitutes arginine to tryptophan at NCF2 and then disrupts the NADPH oxidase complex, nominating *NCF2* as an effector gene **(Fig. 4b)**. The secondary variant rs66977652 (~8kb away from rs13306575, pairwise LD r^2^=0.02) resides in an intron of *NCF2*. Our gene-level analysis showed that rs66977652 confers a significant eQTL effect on *SMG7* (P=3.1×10^-15^ in whole blood; **Fig. 4c** and **Supplementary Figure 3)**, suggesting the secondary signal might influence the risk of SLE by modulating *SMG7* expression, which is supported by functional studies^39^. These findings suggest two potential effector genes, *NCF2* and *SMG7*, in a single locus^25,26^. As another example, a single-variant association test failed to support one effector gene for a novel locus that contains seven protein-coding genes (lead variant rs1 1288784; **Supplementary Figure 2)**. But we found that only *HEATR3* achieved significance in both MAGMA gene-based association analysis and TWAS **(Supplementary Table 10-11)**. *HEATR3* may modulate SLE risk through N0D2-mediated NF-κB signaling^40^. Altogether, the 677 genes implicate roles for cytokine production and signaling, immune responses to stimuli, and the phosphorus metabolic pathway in SLE pathogenesis **(Supplementary Table 12)**, and might inform repurposing drugs approved for musculoskeletal system disorders **(Supplementary Table 13)**.

**Fig. 4.**
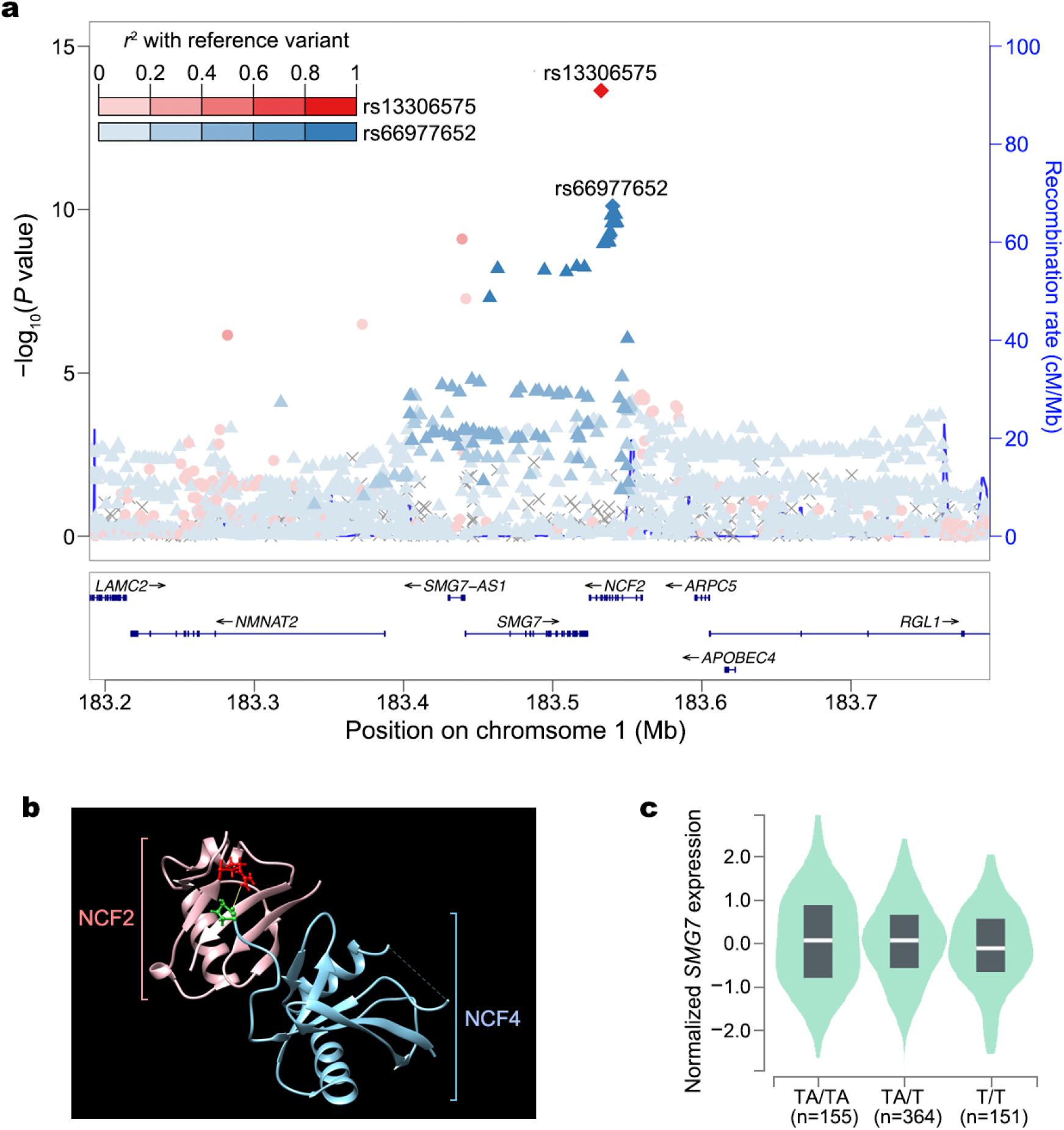
Distinct effects of the two independent association signals (rsl3306575 and rs66977652) at a single locus containing *NCF2* and *SMG7*. **a**, Regional association in the locus including *NCF2* and *SMG7*. Lead and secondary index variants are indicated by diamonds. The lead variant and its LD proxies are in red, while the secondary signal index variant and its LD proxies are in blue. LD was estimated using data from 7,021 unrelated Chinese reference samples, **b**, Arginine at position 395 (red) of NCF2 (pink) is known to interact with proline at position 339 (green) of NCF4 (cyan) via a hydrogen bond (yellow) to form an NADPH oxidase complex. The substitution of arginine at position 395 to tryptophan, induced by the risk allele (A) of the lead missense variant rsl3306575 (p.Arg395Trp) in NCF2, likely disrupts this interaction, **c**, The protective T allele of the secondary signal rs66977652 is associated with decreased expression levels of *SMG7* in a wide range of GTEx v8 human tissues, including whole blood (P=3.1×10^-15^). Sample sizes according to rs66977652 genotypes are included in parentheses on the x-axis. The white line in the center of each box indicates median expression value, while the box for each genotype represents the interquartile range of *SMG7* expressions.

To assess the proportion of phenotypic variance explained by common variants, we applied LD score regression^21^ to the meta-analysis results. Assuming a population prevalence of 0.03% for SLE^9^, we estimated the liability-scale SNP-based heritability from all non*-HLA* variants as h^2^sNP0=1217.24% (standard error (SE)=0.78%). The 66 known and 46 novel non*-HLA* loci explained 62.6% (SE=4.9%) and 22.1% (SE=2.6%) of this overall SNP-based heritability, respectively.

To evaluate global enrichment for SLE-associated variants in epigenomic features, we annotated SLE-associated variants with 15 Roadmap chromatin states in various immune cells using Genomic Regulatory Elements and Gwas Overlap algoRithm (GREGOR)^41^. SLE-associated variants were most significantly enriched in transcription-activating chromatin states at transcription start sites (TSSs) and enhancers **(Fig. 5a** and **Supplementary Table 14)**.

**Fig. 5.**
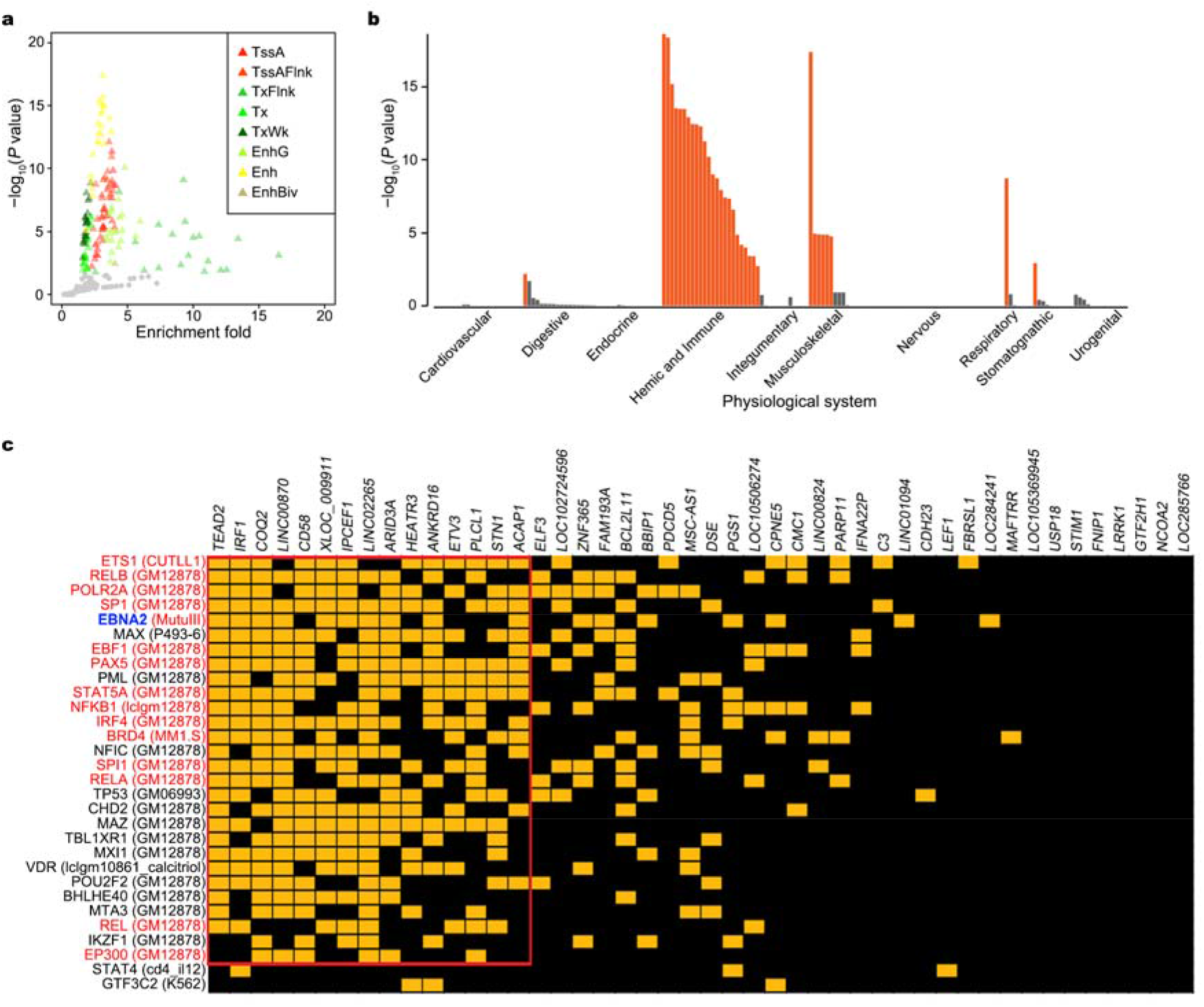
Biological interpretation of SLE associations. **a**, Enrichment of 112 SLE-associated non-HLA variants in ChromHMM core chromatin states across 23 immune cells. X-axis shows fold enrichment while y-axis denotes –log_10_(*P* value of enrichment). Circles and triangles indicate insignificant and significant enrichments, respectively, at FDR<5%. Chromatin states that are significantly enriched for SLE associations are labeled with their ChromHMM annotations: active transcription start site (TssA), flanking active transcription start site (TssAFlnk), transcription at gene 5’ and 3’ (TxFlnk), strong transcription (Tx), weak transcription (TxWk), genic enhancers (EnhG), enhancers (Enh), and bivalent enhancer (EnhBiv). **b**, Physiological systems implicated by the expression of genes within SLE-associated loci (*P*<5×10^-8^). Physiological systems in which the expression of genes from SLE-associated loci are significantly enriched (FDR<5%) are colored orange, **c**, Intersection of 46 novel SLE loci with TF-DNA binding interactions. The x-axis shows SLE loci (named after the nearest genes to the lead variants). The y-axis shows the top 36 TFs, based on probabilities obtained from RELI, sorted in descending order by the number of intersecting loci. Each intersection (yellow box) indicates that the locus (column) contains at least one SLE-associated variant located within a ChIP-seq peak for the given TF (row). The cell type resource for each of the most significant ChIP-seq datasets is indicated in parentheses. TFs that participate in EBNA2 super-enhancers are colored red. EBNA2 protein is highlighted in blue. The red rectangle identifies the optimal cluster of the locus and TF intersections.

To identify tissues and cell types in which SLE-associated genes impact, we tested for expression of SLE-associated genes across human tissues using Data-driven Expression-Prioritized Integration for Complex Traits (DEPICT)^36^. We found significant enrichment for SLE-associated genes not only in the hemic and immune system (P<1.24×10^-3^) but also in musculoskeletal, respiratory, stomatognathic, and digestive tissues **(Fig. 5b, Supplementary Figure 4**, and **Supplementary Table** 15). This might be related with various complication symptoms in multiple organs.

In order to gain further mechanistic insight into non-coding risk variant, we used our RELI method^42^ to identify specific transcription factors (TFs) that significantly occupy SLE risk loci. Eighty-eight TFs concentrate their DNA binding in the 113 SLE loci (relative risk (RR)=2.1-19.0, P<10^-6^; **Supplementary Table 16)**, 40 loci of which (35.4%; including 17 novel loci) were occupied by the EBV Nuclear Antigen 2 (EBNA2) protein (RR=4.9, P=3.95×10^-30^; **Fig. 5c** and **Extended Data Fig. 4)**, which is encoded during the EBV Latency III program, and contributes to B cell transformation. This study achieved a similar finding from independently ascertained SLE cases of East Asian origin, compared with a previous study of Europeans **(Supplementary Table 17)^42^**. In addition, the 46 novel loci were independently associated with EBNA2 (RR=5.0, P<1.7×10^-12^; **Supplementary Table 18)**. Significantly-associated TF Ch1P-seq datasets are enriched for EBV-infected B cell lines, relative to EBV-negative cells (odds ratio (OR)=11.1, P=4.33×10^-55^; **Supplementary Table 19)**. The TFs present at super-enhancers formed upon EBV infection^43^ were remarkably enriched in EBV-positive B cells relative to not EBV-infected cell (OR=22, P=0.001; **Supplementary Table 20-22)**. These results support a role for a gene-environment interaction between SLE loci and EBV infection. We further characterized significant heritability enrichments within 148 cell-state-specific TF profiles for 72 TFs and 123 types of cell or tissue types^44^ (FDR<5%; **Supplementary Table 23)**, which highlight the biological involvement of B cells in SLE pathophysiology.

To identify possible gene regulatory mechanisms impacted by non-coding SLE variants, we performed allelic read imbalance analysis on 985 publicly available ChIP-seq experiments performed in lymphoblast EBV-immortalized B cell lines (see **Online Methods)**. Of the 113 SLE loci, 46 were heterozygous in genotyped cell lines, and 28 exhibited allelic imbalance in at least one ChIP-seq dataset **(Supplementary Table 24)**. The active chromatin histone marks, H3K27ac and H3K4me1, showed strong imbalance in 13 and 8 loci, respectively. For several variants, we observed strong allelic imbalance for particular TFs, suggesting allele-specific function and an underlying genetic mechanism. For example, we observed allelic imbalance at rs2205960 near *TNFSF4*, with EP300, MLLT1, ARID3A, NFATC3, BACH1, MTA2, IKZF1, and BATF all preferring the “G” reference allele, and RUNX3, SMARCA5, TCF12, JUNB, NBN, SKIL, and POU2F2, all preferring the “T” non-reference allele **(Extended Data Fig. 5)**.

To explore cell type-specific regulatory mechanism for non-coding variants, we applied a machine learning approach to predict variants’ mutation effects in 347 types of primary cells and tissues^45^. We identified 71 variant-gene-cell sets showing robust mutation effects on gene transcription in immune cells **(Supplementary Table 25)**. The alternative alleles of rs77571059 and rs200489061 have mutation effects on up-and down-regulating *IRF5 and BLK*expressions, respectively **(Extended Data Fig. 6)**, both of which are consistent with previous *in vitro* experiments^46,47^.

To explore shared genetics between SLE and various traits, we calculated genetic correlations of SLE with 39 complex diseases and 59 quantitative traits in Biobank Japan participants using bivariate LD score regression^48^ **(Supplementary Table 26)**. As expected, we detected significant positive genetic correlations between SLE and two other autoimmune diseases: rheumatoid arthritis (r_g_=0.437) and Graves’ disease (r_g_=0.318). In addition, we found unreported genetic correlations (FDR<0.05) with albumin/globulin ratio (r_g_=-0.242) and non-albumin protein (r_g_=0.238). These findings may reflect the renal complications in SLE patients who have been reported to have significantly lower albumin/globulin ratio and higher serum globulin than healthy controls in epidemiological studies^49^.

We performed the largest GWMA for SLE in East Asians, reiterating the benefits of investigating genetic predispositions to SLE in less-studied populations. We illustrate the advantages of using genetic data to improve knowledge of disease genes, effector tissues, regulatory mechanisms, and biological pathways involved in SLE etiology. These findings elucidate many aspects of SLE genetics and biology and have implications for precision health in SLE.

## Online Methods

### Genome-wide association analyses in eight SLE case-control data sets

We newly recruited 10,029 SLE cases and 180,167 controls in three independent sets and genotyped them on the Illumina Infinium OmniExpress Exome-8 Array, Illumina Infinium Global Screening Array, or Korean Biobank Arrays^50^ **(Supplementary Table 1)**. Cases were diagnosed by medical specialists using American College of Rheumatology classification criteria for SLE^51^. Controls had neither SLE nor family history of SLE. Written informed consent was obtained from all participants. Protocols were approved by institutional review boards in participating institutions.

To improve statistical power for discovering genetic effects, we revisited raw genome-wide genotypes from five published studies^1,2,4,5,8^ **(Supplementary Table** 1). Quality controls were then conducted for each of the eight data sets. Briefly, we excluded individuals of: 1. call rate<95%; 2. mismatch between ascertained and genotype-inferred sex; 3. outliers for heterozygosity rate; 4. population outliers from the East Asian cluster in principal component analysis (PCA) of genotypes against 1000 Genomes Project (1KGP) populations^19^. For quality controls of genetic variants, we excluded variants with any of the following criteria: 1. call rate<99% for Japanese data sets or <95% for the remainder; 2. P value for Hardy-Weinberg equilibrium (P_HWE_)<1.0×10^-6^ in the controls; 3. minor allele counts (MAC)<10 for Japanese data sets and MAF≤1% for the others. We then conducted genotype imputation for each data set separately. Haplotypes were estimated using SHAPEIT^52^ or Eagle 2^53^. Genotype imputation was accomplished using reference panels from the 1KGP phase 3 v5^19^ and IMPUTE2/4^54,55^, or MINIMAC4^56^. For genotype imputation in individuals from Korea and Japan, we additionally used population-specific reference panels from 397 Korean Reference Genome Project^20^ and 7,472 whole-genome sequencing datasets, respectively.

We tested association between SLE risk and genotype dosages in each data set using a logistic regression or linear mixed model in PLINK^57^, SNPTEST^58^, or EPACTS (https://genome.sph.umich.edu/wiki/EPACTS) **(Supplementary Table 1)**. Within each data set, we filtered out association results based on imputation quality (IMPUTE info or MINIMAC r^2^≤0.3), MAF≤0.5%, or P_HWE_<1.0×10^-6^. For each cohort, the association analysis for the X chromosome was conducted separately by sex and then meta-analyzed across both men and women. For data sets analyzed using a linear mixed model **(Supplementary Table 1)**, allelic effects and standard errors were converted to a log-odds scale to correct for case-control imbalance^59^.

### Fixed-effects meta-analysis

We aggregated the association summary statistics from the eight data sets using a fixed-effects inverse-variance meta-analysis in METAL^60^. We applied a genomic control correction to each association summary statistic. Heterogeneity in allelic effect sizes among data sets was assessed using Cochran’s Q statistic. We excluded genetic variants available in only a single data set. We defined SLE susceptibility loci by merging ±250 kb windows around genome-wide associated variants to ensure that lead SNPs were at least 500 kb apart. We defined lead variants as the most significant SLE-associated variant within each locus. A locus was considered novel if the lead SNP was at least 500 kb away from any previously reported SLE-associated variants.

### Approximate conditional association analysis

To dissect distinct association signals at each SLE locus, we performed an approximate conditional analysis using GCTA COJO^22^ with genome-wide meta-analysis summary statistics based on LD estimated from 7,021 unrelated Chinese controls. The Chinese reference individuals for LD calculation were retrieved from the Chinese study using the Illumina Infinium Global Screening Array data **(Supplementary Table 1)**, excluding first-and second-degree relatives. We excluded genetic variants that have substantial MAF differences (>0.05) between meta-analysis summary statistics and the reference individuals. We only presented signals below a conditional threshold of P<5×10^-8^.

### Bayesian statistical fine-mapping analysis

To prioritize causal variants in SLE susceptibility loci, a statistical fine-mapping analysis was performed using FINEMAP v1.4 software^29^, with meta-analysis z-scores and LD matrices estimated from the 7,021 Chinese reference individuals. We used default priors and parameters in FINEMAP, assuming at most five causal signals in the ±250 kb region around a lead variant at each SLE locus, excluding the *HLA* region (chromosome 6: 25-34 Megabases (Mb) in build hg19) and 7qll.23. FINEMAP was used to estimate the PPs and Bayes factor values of potential causal configurations and variant-level PPs for causality calculated for individual variants. We then built the 95% posterior credible sets of causal variants. The causal configurations were prioritized based on configuration-level PPs.

### Assay for transposase-accessible chromatin using sequencing (ATAC-seq) in blood CD4^+^ T and CD19^+^ B cells

We collected fresh whole blood samples from five healthy volunteers of Chinese population. CD4^+^ T and CD19^+^ B cells were sorted using fluorescence activated cell sorting and used to create ATAC-seq libraries as previously described^61^. Libraries were sequenced on the BGISEQ 500 platform, with a 50 bp paired-end read (unpublished data) and ATAC-seq peaks were called using MACS2^62^, to detect open accessible elements at the *ACAP1* locus in CD4^+^ T and CD19^+^ B cells. Each participant provided written consent. The study protocol was approved by the institutional review board at the Institution of Dermatology, Anhui Medical University.

### Luciferase Reporter Assay

Three identical copies of the 24 bp element flanking each allele of rs61759532 (5’-TGC TCT GGG G<u>C</u>G GTT AGC AAC TTC-3’ for the *C* allele and 5’-TGC TCT GGG G<u>T</u>G GTT AGC AAC TTC-3’ for the *T* allele) were subcloned into the luciferase vector, pGL4.26 (luc2/minP/Hydro), between the XhoI and BglII sites upstream of the minimal promoter for the firefly luciferase gene, to test the enhancing activity of the inserts **(Supplementary Figure 5)**. The firefly luciferase vector (1 μg) and normalizing Renilla luciferase vector (500 ng) were co-transfected into THP1 cells for 2 days using Lipofectamine 3000 (Thermo-Fisher Scientific). Luciferase activity was measured in five independent biological replicates using the Dual-Luciferase Reporter Assay Kit (Promega) according to the manufacturer’s instructions. Relative fold-change in firefly luciferase activity was normalized by both transfection efficiency, based on Renilla luciferase activity, and minimal luciferase activity from the pGL4.26 vector without inserts.

### EMSA

EBV-transformed B or THP1 cells were grown in RPMI1640 medium including 10% fetal bovine serum and 1% penicillin/streptomycin. EMSA probes were constructed by annealing biotin-conjugated 30-residue oligonucleotide sequences flanking rs61753158: 5’-biotin-ACC TGC TCT GGG G<u>C</u>G GTT AGC AAC TTC CTG-3’ (forward) and 5’-biotin-CAG GAA GTT GCT AAC C<u>G</u>C CCC AGA GCA GGT-3’ (reverse) for the *C* allele; 5’-biotin-ACC TGC TCT GGG G<u>T</u>G GTT AGC AAC TTC CTG-3’ (forward) and 5’-biotin-CAG GAA GTT GCT AAC C<u>A</u>C CCC AGA GCA GGT-3’ (reverse) for the *T* allele. EMSA was performed using the LightShift Chemiluminescent EMSA Kit (Thermo-Fisher Scientific) according to the manufacturer’s instructions. Briefly, a nuclear extract (10 μg) of EBV-transformed B or THP1 cells was incubated with EMSA probes (20 fmol) for 30 min at room temperature in a final volume of 15μl with lx EMSA binding buffer, after pre-incubation with a non-specific competitor poly(dI-dC) and 0 or 4 pmol of a specific, non-conjugated competitor. DNA-protein complexes were separated on 6% nondenaturing polyacrylamide gel.

### DEPICT analysis

We used DEPICT vl release 19 4^36^ to prioritize genes and tissues and cells implicated by our genome-wide association meta-analysis results. All of the genetic variants with P<5×10^-8^ were included. Input variants were clumped in DEPICT, using default 500 kb flanking regions with an LD cutoff of r^2^>0.1, based on 1KGP East Asians data, yielding 1,521 autosomal loci. We applied a threshold of false discovery rate (FDR)≤0.01 to declare significant gene and tissue/cell enrichment.

### TWAS analysis

We performed a transcriptome-wide association analysis using meta-analysis summary statistics in FUSION^33^ to infer gene expression changes in SLE. The training data sets for imputing gene expression were generated using eQTLs from 105 Japanese individuals in six different cell types: B cells, CD4^+^ T cells, CD8^+^ T cells, monocytes, natural killer (NK) cells, and peripheral blood cells^34^. We defined a significance threshold at Benjamini-Hochberg FDR of 0.05 to correct for multiple testing of each cell type.

### Gene mapping using Functional Mapping and Annotation of Genome-Wide Association Studies (FUMA)

Potentially disease-causing genes at each SLE locus were mapped using FUMA vl.3.6^37^ with three mapping strategies: physical, eQTL, and chromatin interaction mapping. Briefly, genes within a 10 kb window from lead and proxy variants (r^2^≥0.6 in the 1KGP East Asians) were selected by physical mapping. eQTL mapping identified genes that were potentially cis-regulated by SLE variants within ≤1 Mb distance from lead variants, using known eQTL variants with FDR-corrected P values<0.05 in immune-related cell data in eQTL databases **(Supplementary Table 27)**. Variants were strictly filtered using combined annotation dependent depletion (CADD) scores^63^ (≥12.37), maximum RegulomeDB scores^64^ (≤7), and maximum 15-core chromatin state^65^ (≤7; open chromatin) in any blood cell type, and FANTOM5^66,67^ promoter and enhancer regions. Chromatin interaction mapping was used to search for genes whose promoters had chromatin interactions with cell type-specific enhancers containing SLE variants (CADD score ≥ 12.37, RegulomeDB score ≤ 7), using Hi-C data from EBV-transformed B cells and spleen (GSE87112)^68^, and annotation of promoter and enhancer regions in various blood cell types and spleen^69^.

### MAGMA-based gene prioritization

MAGMA v1.0 7^35^ was deployed to calculate gene-level disease association P values from variant-level association summary statistics within genes using a variant-wide mean model with data from the 1KGP East Asian reference panel.

### Heritability estimation by LD score regression

Overall SLE heritability *h*^2^ explained by genome-wide variants was estimated using the LD score regression model^21^ with LD scores^19^ from 1KGP East Asian descendants, based on an SLE population prevalence of 0.03% in East Asian populations^9^. SLE heritability estimate was further partitioned according to known and novel SLE loci and transcription factor binding sites (TFBSs), to assess whether variants within the selected annotations explained significantly more SLE heritability using stratified LD score regression^70^. The boundary of each SLE locus was arbitrarily defined as ±500 kb flanking a lead SLE-risk variant. TFBS annotations were obtained from IMPACT^44^. IMPACT provides 707 tissue-specific TFBSs annotation sets for 137 TFs in 23 tissues, including 58 sub-cell types. Heritability enrichment estimates for query annotations *i* (*E_i_*) were calculated as follows:

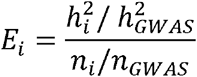

where 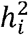 is heritability explained by variants within the query annotation *i*, 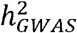 is the overall SLE heritability attributable to genome-wide variants, *n_i_* is the number of variants in the query annotation *i*, and *n_GWAS_* is the total number of variants analyzed. Standard errors were estimated using the block jackknife method^70^. P-values were calculated based on the Z score and corrected by an FDR of 0.05. *HLA* variants were excluded from the TFBS-based partitioned heritability enrichment analysis.

### Enrichment analysis for epigenomic features using GREGOR

Enrichment of SLE-associated variants in epigenomic regulatory features was evaluated using GREGOR^41^. We utilized the Roadmap ChromHMM annotation comprising 15 chromatin states inferred by a combination of multiple histone marks in 23 immune cell types f https://egg2.wustl.edu/roadmap/web_portal/). GREGOR generated 1,000 random lead variant sets with three properties for the actual SLE-risk lead variant set (distance to the nearest gene, MAF, and number of proxy variants in LD at an *r^2^* threshold of 0.8 in the 1KGP East Asian populations), to find a null distribution for the number of random lead variants in the annotation of interest, accounting for proxy variants. Enrichment P values were derived from the upper tail for the number of actual lead SLE-risk variants in the query annotation in the null distribution, using saddle point approximation and FDR correction at 5%.

### Regulatory Element Locus Intersection (RELI) analysis

The RELI algorithm^42^ was used to estimate the significance of intersections between the genomic coordinates of SLE loci (defined as the lead variants and their strong LD proxies with r^2^>0.8 in 1KGP East Asians) and the DNA sequences bound by a particular TF or co-factor, as determined by ChIP-seq. We used 1,544 ChIP-seq datasets as previously described^42^, which contains 1,536 ChIP-seq datasets for 344 human TFs in 221 cell lines and eight viral ChIP-seq datasets from EBV-infected B cells for the EBV gene products EBNA1, EBNA2 (three datasets), EBNA3C, EBNA-LP, and Zta and from HIV-infected T-cells for TAT. We identified 371 ChIP-seq datasets for TFs from various cell types, which had been previously suggested to form super-enhancers upon EBV infection in EBV-infected and transformed B cells^43^. Using RELI, we computed the significance and enrichment level for each ChIP-seq dataset by comparing the observed intersections with a null distribution of intersections obtained from 2,000 simulations. For each simulation, genetic variants were randomly chosen throughout the genome, ensuring their MAFs and LD structures similar to the actual lead variants and LD proxies. We used a significance threshold of P < 10^-6^ after Bonferroni correction and computed relative risk by dividing the observed intersections by the mean expected number of intersections.

### Identification and processing of public B cell line chromatin immunoprécipitation and sequencing (ChIP-seq) datasets for allelic analysis

We identified 1,078 ChIP-seq datasets from experiments performed in B cell lines in the Gene Expression Omnibus (GEO)^71^ using custom text searching scripts. 505 datasets were obtained for the GM12878 cell line and 573 for non-GM12878 cell lines. Annotations were manually checked for every dataset (assay type, cell line, and assayed molecule) to ensure accuracy. Sequence Read Archive (SRA) files obtained from GEO, representing sequencing reads, were analyzed using an automated pipeline. Briefly, the pipeline first ran quality control (QC) on the FastQ files using FastQC (v0.11.8)^72^. If FastQC detected adapter sequences, the pipeline ran the FastQ files through Trim Galore (v0.4.2)^73^, a wrapper script that runs cutadapt (v1.9.1)^74^, to remove the detected adapter sequences from the reads. The quality controlled reads were then aligned to the reference human genome (hg19/GRCh37) using bowtie2 (v2.3.4.1)^75^, followed by sorting using samtools (v1.8.0)^76^ and removing duplicate reads using picard (v1.89)^77^. Finally, peaks were called using MACS2 (v2.1.2)^62^, with four “modes” using the following parameter settings: M0DE1 = -g hs -q 0.01; MODE2 = -g hs -q 0.01 -broad; MODE3 = -g hs -q 0.01 --broad --nomodel -extsize 500; and M0DE4 = -g hs -q 0.01 -broad -nomodel - extsize 1000. Peaks were merged across the four MODES using bedTools to produce a final peak set for each experiment. ENCODE blacklist regions^78^ were removed from the peak sets using the hg19-blacklist.v2.bed.gz file available at https://github.com/Bovle-Lab/Blacklist/blob/master/lists/hg19-blacklist.v2.bed.gz. 93 datasets failed at the download, alignment, or peak calling steps, yielding a total of 985 ChIP-seq peak sets in .BED format for subsequent analysis.

### Allele-dependent ChIP-seq data analysis

We used whole genome sequencing data from 1KGP phase 3 to identify heterozygous genetic variants in B cell lines. Starting with variant call files produced by the 1KGP, heterozygous variants were identified for each subject. For the GM12878 cell line, we used genotyping information obtained from Illumina OMNI-5 arrays. Genotypes were called using the Gentrain2 algorithm within Illumina Genome Studio. Quality control was performed as previously described^79^. Quality control data cleaning was performed in the context of a larger batch of non-disease controls to allow for data quality assessment. Briefly, all cell lines had call rates >99%; only variants with MAF≥0.01 were included; and all variants with PHWE>10^-4^ were included. We performed genome-wide imputation using overlapping 150 kb sections of the genome with IMPUTE2^55^ and 1KGP phase 3 (June 2014), and then filtered imputed variants with probability<0.9 or imputation quality<0.5 in addition to the same criteria described above for typed markers. Regions of the genome with abnormal chromosome counts (i.e., regions that did not have two chromosomes) were removed from consideration using the cnvPartitionsoftware package (Illumina Genome Studio) with default parameter settings, due to their potential effects on allelic read imbalances.

Next, as a final quality control step, the identity of cell lines, as annotated in GEO, was confirmed by assessing read counts at variants identified as heterozygous. For each dataset in each cell line, we examined all heterozygotes with at least five sequencing reads. We then calculated the fraction of these variants with exactly zero reads on the weak allele (i.e., the allele with fewer mapped sequencing reads). Zero weak allele reads is a hallmark of a mis-annotated cell line, since a variant that is thought to be a heterozygote but in reality is a homozygote will always exhibit zero weak allele reads. Any dataset ≥ 45% “zero weak allele read heterozygotes” was flagged as a mis-annotation by the original producers of the dataset and removed from downstream analyses. This cutoff was chosen based on comparisons between purposely matched and mis-matched genotyping array/ChIP-seq experiment pairs. In total, 482 datasets were used for allelic analyses.

To identify possible mechanisms underlying identified allelic variants, we applied the MARIO method^42^ to the ChIP-seq dataset collection. Briefly, MARIO identifies common genetic variants that are (1) heterozygous in the assayed cell line and (2) located within a peak in a given ChIP-seq dataset. It then examines the sequencing reads that map to each heterozygote in each peak for imbalance between the two alleles. MARIO Allelic Reproducibility Score (ARS) values >0.4 were considered allelic, following our previous study^42^. We also used an additional filter to ensure consistency across ChIP-seq datasets. Specifically, for each variant/regulatory molecule pair, we calculated the fraction of ChIP-seq datasets that shared the same preferred allele. For example, if a particular variant was a heterozygote located inside an H3K27ac ChIP-seq peak in 10 datasets, and five of them preferred the G allele (with ARS value > 0.4), this value would be 0.5. We only included variant/regulatory molecule pairs where this value was ≥ 0.3.

### In-silico mutagenesis to pinpoint causal variants and genes

To explore the mutation effect and then to prioritize the causal variants and potential regulatory mechanisms for SLE genetic associations, we applied Mutation effect prediction on ncRNA transcription (MENTR)^45^ analysis in significant summary-level associations from current meta-analysis. MENTR learns cell-type-specific transcription probability of promoters and enhancers surrounding transcription start sites (TSSs) using FANT0M5 Cap Analysis of Gene Expression (CAGE)^80^ from 347 types of samples comprising a variety of primary cells and tissues, and then predicts mutation effects on transcripts by *in silico* mutagenesis for certain variants based on the degree of probability change. In order to investigate mutation effects for 9,506 significant non*-HLA* SNPs (P<5×10^-8^) on transcription, we searched for genes whose TSS are less than 100 kb from any significant non*-HLA* SNP and found 350,410 variant-gene pairs. We then evaluated the mutation effect for these 350,410 variant-gene pairs in MENTR. We identified 2,383 top variant-gene-cell sets that had robust mutation effects at mutation effect>0.1, which shows high accuracy of prediction^45^. Among these top variant-gene-cell sets, we eventually restricted the results to the 71 sets that contain lead variants, proxies of strong LD with lead variants (r^2^>0.8 in the combined genotypes of 1KGP phase 3 v5 East Asians and whole genome sequencing data for 7,472 Japanese unrelated individual), and immune cells.

### Genetic correlation between SLE and diseases/traits using LD score regression

We calculated genetic correlations between 98 traits (39 diseases^17^ and 59 quantitative traits^81^) and SLE by using LD score regression^48^. SNPs were restricted to HapMap3, because these are well-imputed in most studies. The *HLA* region was excluded because of its complex LD structure. LD scores of the 1KGP East Asians were used to estimate genetic correlations. Significance was defined as Benjamini-Hochberg FDR of 0.05.

## Data Availability

The ATAC-seq data for human blood B and T cells have been deposited with the China National Genomics Data Center (https://bigd.big.ac.cn/gsa-human/browse) under accession no. HRA000271. The expression quantitative trait loci summary-level data in blood immune cells are publicly available from our website (JENGER; http://jenger.riken.jp/en/). The meta-analysis summary-level results will be publicly available following 6 month embargo from the date of publication. All other data supporting the findings of this study are available within the paper and its supplementary information files and from the corresponding author upon reasonable request.

## Acknowledgements

We acknowledged the participants in this study. We appreciate the contribution of Japanese Research Committee on Idiopathic Osteonecrosis of the Femoral Head. We appreciate all contributors to BioBank Japan. Details are included in supplementary material. This research was supported by General Program (81872516, 81573033, 81872527, 81830019, 81421001), Young Program (81803117), Exchange Program (81881340424), and Science Fund for Creative Research Groups (31630021) of National Natural Science Foundation of China (NSFC), Distinguished Young Scholar of Provincial Natural Science Foundation of Anhui (1808085J08), National Program on Key Basic Research Project of China (973 Program) (2014CB541901), Science Foundation of Ministry of Education of China (213018A), Program for New Century Excellent Talents in University of Ministry of Education of China (NCET-12-0600), The Bio & Medical Technology Development Program of the National Research Foundation, funded by the Ministry of Science & ICT of the Republic of Korea (NRF-2017M3A9B4050355 to S.C.B.), Basic Science Research Program through the National Research Foundation of Korea funded by the Ministry of Science, ICT and Future Planning (2015R1C1A1A02036527 and 2017R1E1A1A01076388 to K. Kim), National BioBank of Korea, the Centers for Disease Control and Prevention, Republic of Korea (KBN-2018-031 to S.S.L.), Center for Genome Science, Korea National Institute of Health, Republic of Korea (4845-301, 3000-3031 to M.Y.H., K. Yoon and B.J.K.), Japan Agency for Medical Research and Development (AMED) and the BioBank Japan project supported by the Ministry of Education, Culture, Sports, Sciences and Technology of the Japanese Government and AMED under grant numbers (17km0305002 and 18km0605001), Grant of Japan Orthopaedics and Traumatology Research Foundation, Inc, (No.350 to Y.Sakamoto), RIKEN Junior Research Associate Program (to H.S.), US NIH grants (AI024717, AI130830, AI148276, HG172111 and AR070549 to J.B.H.), US Department of Veterans Affairs (BX001834 to J.B.H.), and Center for Pediatric Genomics Award and CCRF Endowed Scholar Award of Cincinnati Children’s Hospital (to M.T.W.). We appreciate the generous gift of pGL4.26 vector from Prof. Joon Kim at Graduate School of Medical Science Engineering, KAIST, Daejeon, South Korea.

## Author Contributions

X.Y., K. Kim and H.S. contributed equally to this work, and either has the right to list himself first in bibliographic documents. S.C.B., Y.C., C.T., X. Zhang, X.Y., K. Kim and H. S. conceived the study design. S.C.B., Y.C, X. Zhang, S.Y., K. Kim and C.T. acquainted the financial support. X.Y., K. Kim, H.S., C.T., Y.C. and S.C.B. wrote the manuscript. X.Y., K. Kim, H.S., E.H., X. Zheng, V.L. and Y.W. conducted all of the analyses with the help of J.B.H., L.C.K., M.T.W., S.P., S.E., H.S., K.T., N.O., M.K., K.I. and C.Terao. K. Kim, S.Y.B, L.W., L.L., R.X.L., Y. Sheng, M.Y.H., W.L., K. Yoon, M.C., H.H., M.W., Y. Tang, H.D., C.L., C.S., W.F., K.L., B.J.K., H.S.L., S.C.B., S.H., Y.Sakamoto, N.Sugano, M.M., D.T., K.Karino, T.Miyamura, J.N., G.M., T.Kuroda, H.N., T.Miyamoto, T.T., Y.Kawaguchi, K.A., Y.Tada, K.Yamaji, M.S., T.A., A.S., T.Sumida, Y.Okada, K.Matsuda, K.Matsuo, Y.Kochi, T.Seki, Y.Tanaka, T.Kubo, R.H., T.Yoshioka, M.Y., T.Kabata, Y.A., Y.Ohta, T.O., Y.N., A.K., Y.Y., K.Ohzono, K.Yamamoto, K.Ohmura, T.Yamamoto and S.I. generated genetic data. L.L. and H.H. contributed to ATAC-seq experiment. S.J. and T.H.K. performed luciferase reporter assays and EMSAs. S.Y.B., Y.C.K., W.T.C., S.S.L., S.C.S., Y.M.K., D.Y., C.H.S., Y.B.P., J.Y.C., Y.P., G.YA., J.M.S., Y.K.L., W.Y., S.Y., B.J.K., N. Shen, H.S.L., X. Zhang, C.T. and S.C.B. managed the cohort data.

## Competing Interests

The authors declare no competing interests.

---

**Extended Data Fig. 1.**
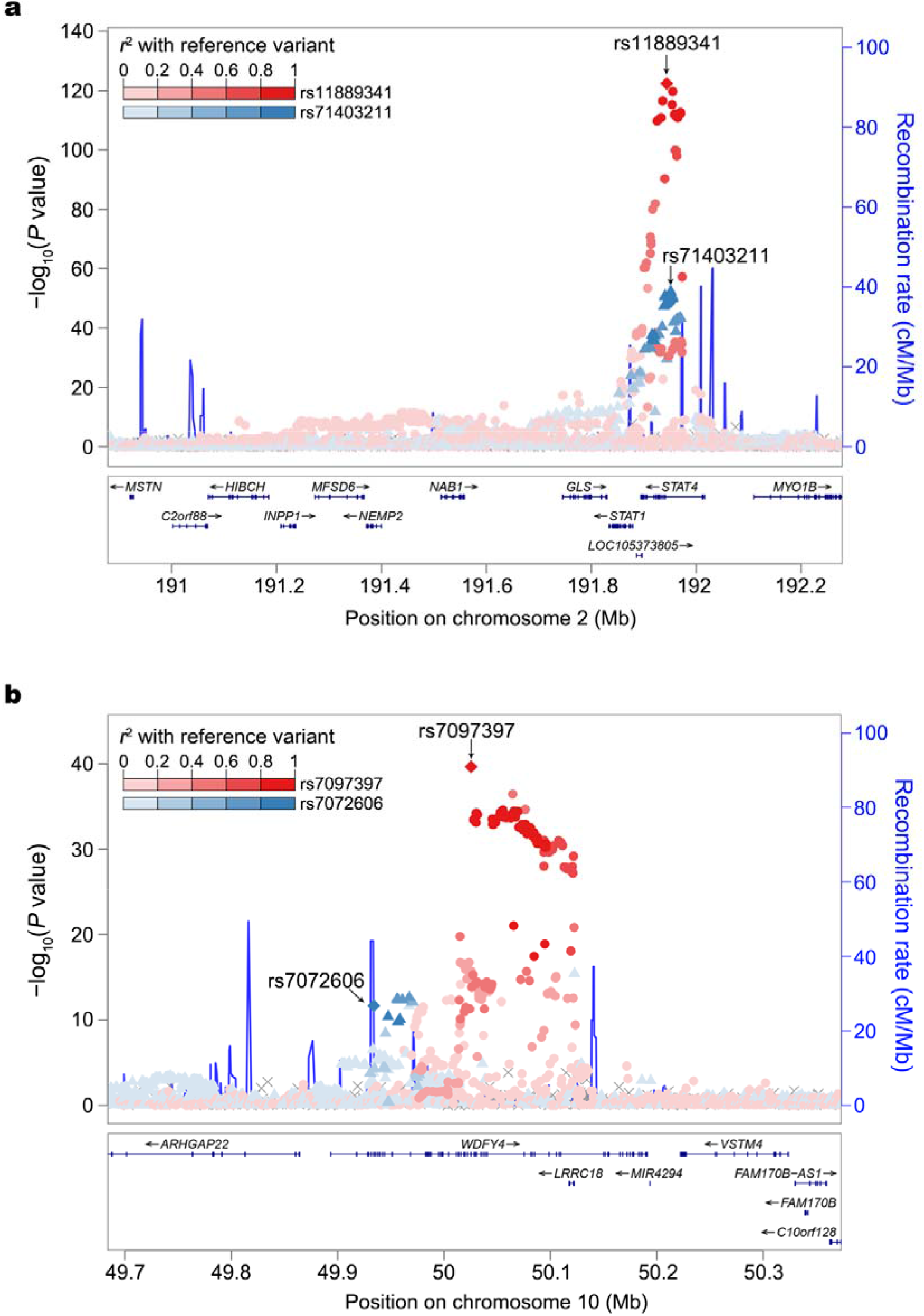
Two independent association signals identified. **a**, at two intronic variants within known *STAT4* locus. **b**, at known (rs7097397, p.Arg1816Gln) and new (rs7072606, p.Ser214Pro) missense variants within *WDFY4* locus. The lead and secondary index variants are labeled in diamond. The lead variant and its LD proxies are in red while the secondary signal index variant and its LD proxies are in blue. The LD is estimated from 7,021 Chinese samples.

**Extended Data Fig. 2.**
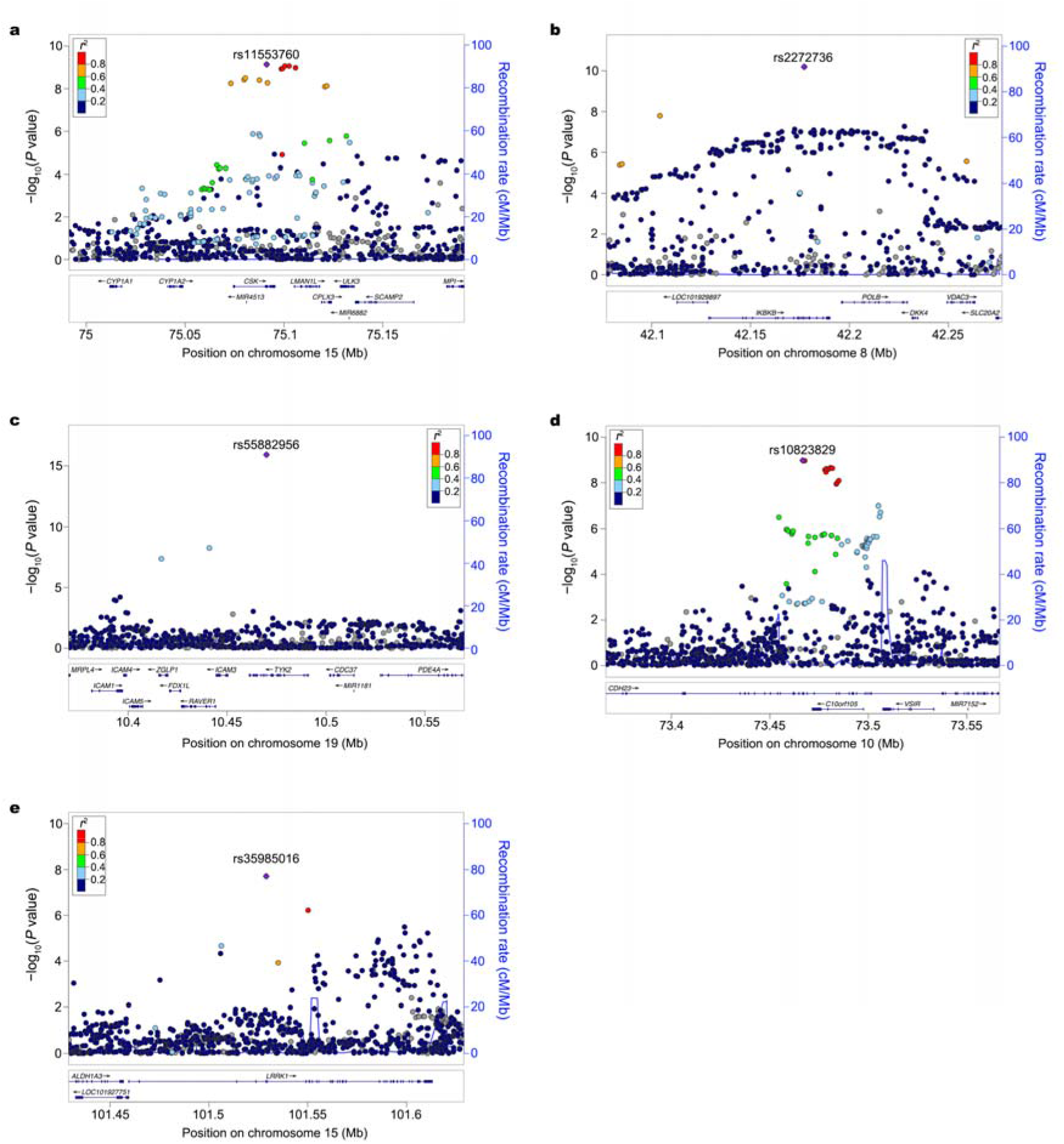
New lead exonic variants identified at three known *(CSK IKBKB*, and *TYKZ)* and two novel (*CHD23* and *LRRK1)* loci. **a**, rs11553760 (synonymous variant) at *CSK*. **b**, rs2272736 (p.Arg303Gln, missense variant) at *IKBKB*. **c**, rs55882956 (p.Arg703Trp, missense variant) at *TYK2*. **d**, rs10823829 (synonymous variant) at *CHD23*. **e**, rs35985016 (p.Lys203Glu, missense variant) at*LRRK1*. The lead SNP is labeled as purple diamond. The LD is estimated from 7,021 Chinese samples.

**Extended Data Fig. 3.**
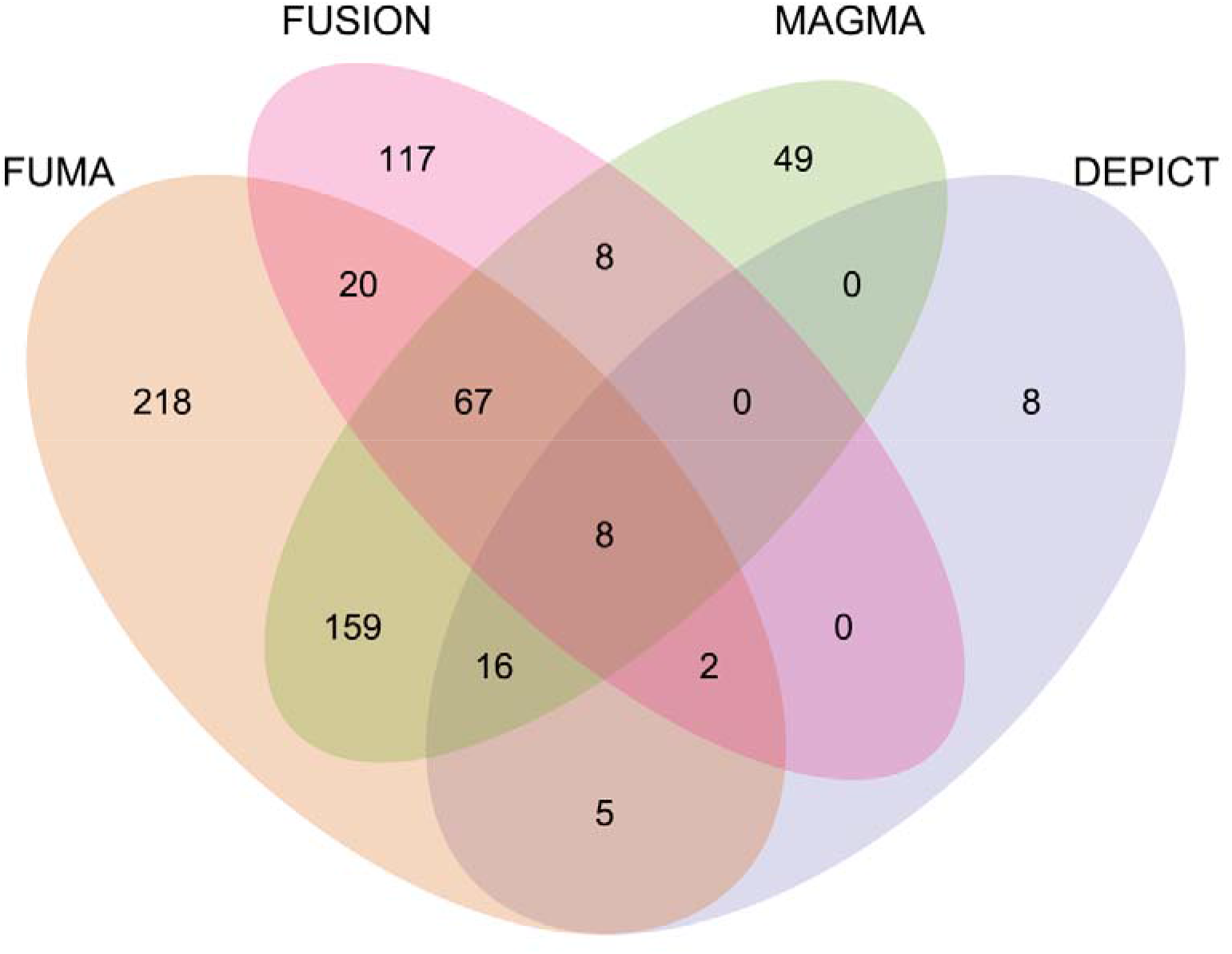
Venn diagram of significant gene findings from four gene-level approaches. The significant gene findings from FUMA, FUSION, MAGMA, and DEPICT are in light orange, light read, light green, and light purple, respectively.

**Extended Data Fig. 4.**
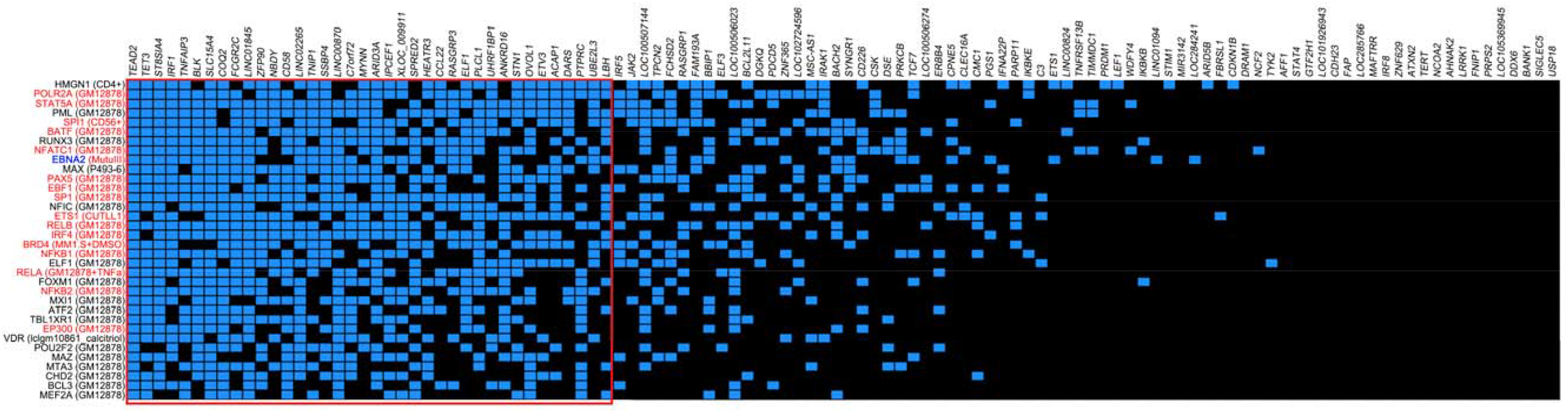
Intersection of 112 non-HLA SLE risk loci with TF-DNA binding interactions with the genome. The x-axis displays SLE loci (named after the nearest genes to the lead variants). The y-axis displays the top 36 TFs, based on probabilities obtained from RELI, sorted in descending order by the number of intersecting loci. Each intersection (yellow box) means that the locus (column) contains at least one SLE-associated variant located within a ChIP-seq peak for the given TF (row). The cell type resource for each of the most significant ChIP-seq dataset is indicated in parentheses. TFs that participate in EBNA2 super-enhancers are colored red. The red rectangle identifies the optimal cluster of the locus and TF intersections.

**Extended Data Fig. 5.**
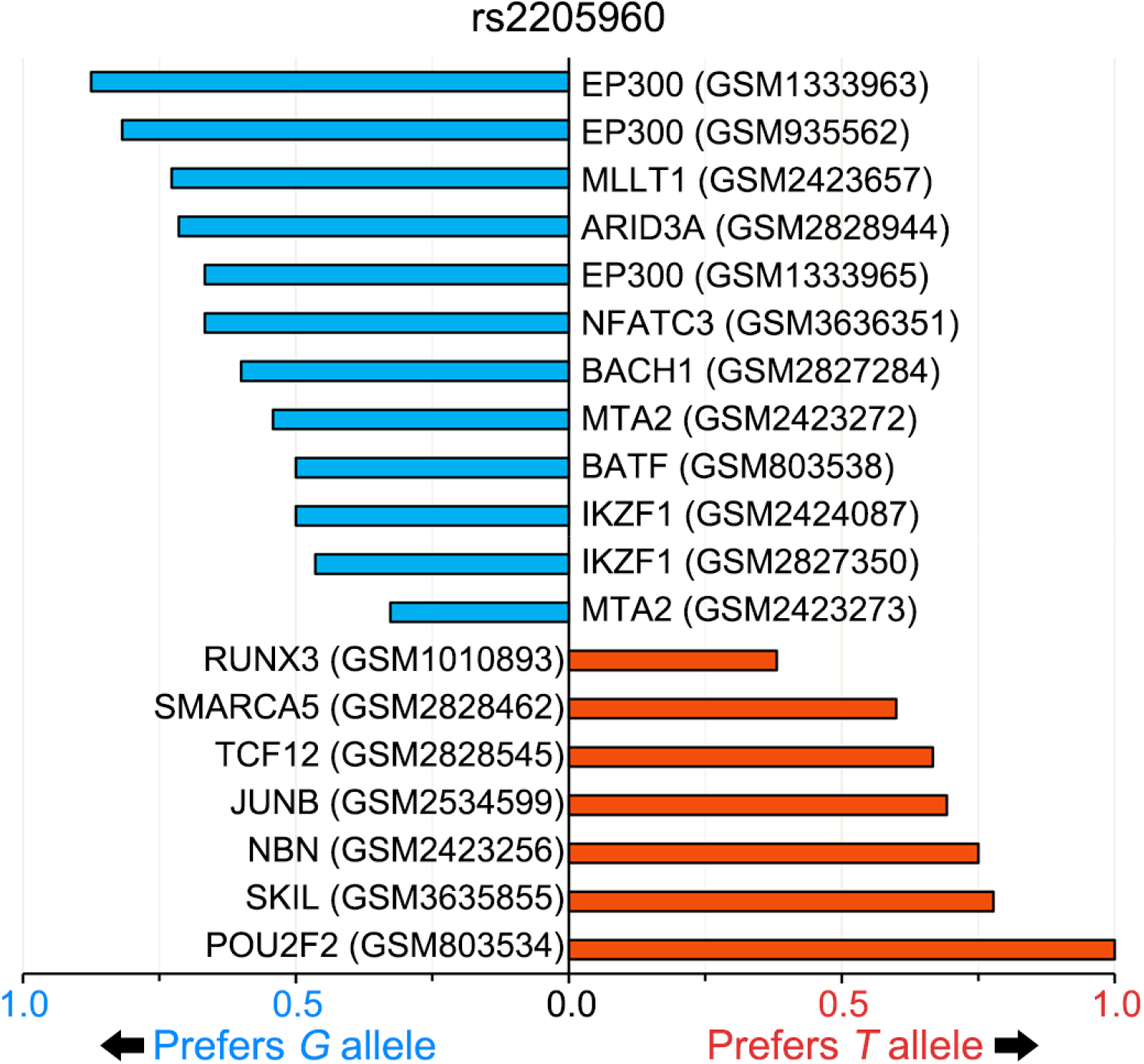
Allele-dependent regulatory proteins binding to the rs2205960 variant. Allele-dependent binding of regulatory proteins at the *TNFSF4* locus based on ChIP-seq read allelic imbalance analysis (see Online Methods). The x-axis indicates the preferred allele, along with a value indicating the strength of the allelic behavior, calculated as 1 minus the ratio of the weak to strong reads (for example, 0.5 indicates the strong allele has approximately twice the reads of the weak allele).

**Extended Data Fig. 6.**
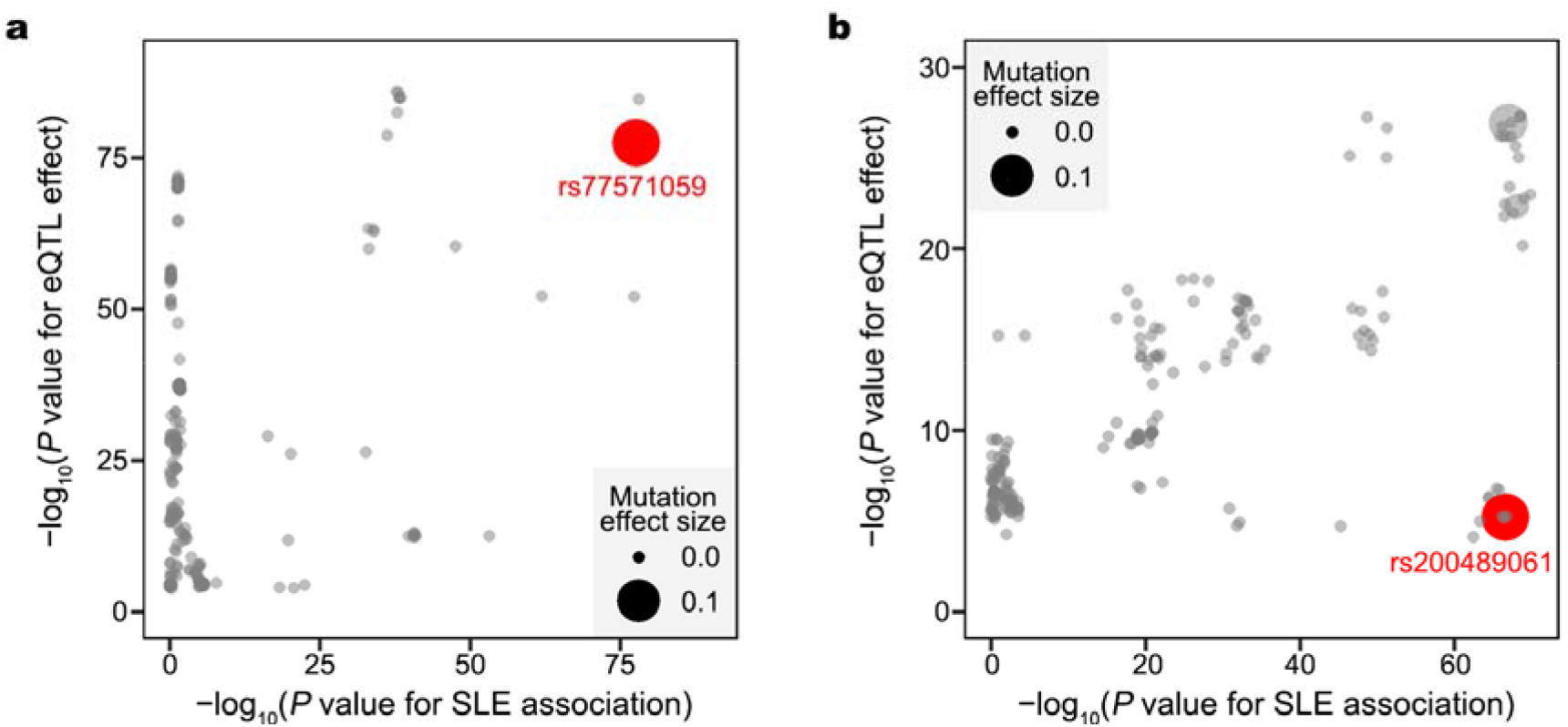
Robust mutation effect predicted in MENTR. **a**, rs77571059 on regulating *IRF5* expression in neutrophil, **b**, rs200489061 on regulating *BLK* expression in natural killer cell. The x-axis is the minus log_10_ transformed single-variant association P values for SLE risk while the y-axis denotes the minus logio transformed eQTL P values in GTEx v8 whole blood. The size of each dot is scaled by the predicted mutation effect magnitude. The variants with robust mutation effect are in red.

## Notes

### Competing Interest Statement

The authors have declared no competing interest.

### Author Declarations

institutional review boards in participating institutions

